# Metabolomic Signatures of Habitual Physical Activity in Childhood

**DOI:** 10.64898/2026.10.30.26366930

**Authors:** Lara Vehovec, Jill Fründ, Jeannie Horak, Kathrin Guerlich, Hans Demmelmair, Mireia Alcázar, Werner Römisch-Margl, Fabian Geyer, Elvira Verduci, Dariusz Gruszfeld, Annick Xhonneux, Joaquín Escribano, Berthold Koletzko, Veronica Luque, Veit Grote, Gabi Kastenmüller

## Abstract

Physical activity (PA) supports health across the life course, yet the biological pathways linking habitual PA to long-term health remain incompletely understood. Metabolomics provides a powerful tool to study PA-related metabolic adaptations, but how habitual PA shapes the metabolome in childhood remains poorly characterized. Addressing this gap, we examined cross-sectional associations between accelerometer-measured habitual PA and the plasma metabolome in 515 children aged 5.5 to 11 years from the European Childhood Obesity Project (CHOP) and additionally determined whether associations differed between sexes and across the body mass index (BMI) range. Using targeted metabolomics, we quantified 315 metabolites and derived 91 metabolite sums and ratios. Associations with PA were assessed using linear mixed-effects models. Replication was performed in an independent pediatric cohort. Among the 406 metabolite variables, 73 were associated (p < 0.05) with at least one PA variable, of which 12 replicated, including lysophosphatidylcholine (LPC) a C14:0 and LPCs/phosphatidylcholines (PCs) with higher levels in more active children, and PC aa C40:6, monounsaturated PCs/saturated PCs, and valine/LPC a C18:1 with lower levels in more active children. At the metabolite-class level, intensity-dependent association patterns emerged. Amino acids and acylcarnitines were lower in more active children predominantly for time spent in higher-intensity PA, whereas LPCs were higher mainly with more total PA (80% light PA) and less sedentary time. Associations were more pronounced at older ages and differed (p < 0.05) by BMI for 42 metabolite variables and by sex for 45 variables in the interaction analyses. These findings provide insights into PA-related metabolic adaptations early in life and may help elucidate biological pathways linking childhood PA to long-term health.

## Introduction

Lack of physical activity (PA) is one of the leading risk factors for many non-communicable diseases and chronic conditions such as insulin resistance^1–3^, metabolic syndrome^4,5^, and cardiometabolic diseases^6–8^ across the entire lifespan. Various metabolic mechanisms have been implicated in conveying the health benefits of PA. These include mitochondrial biogenesis and adaptations^9^, improved insulin sensitivity through exercise-induced 5’ adenosine monophosphate-activated protein kinase (AMPK) activation that promotes glucose transporter type 4 (GLUT4) translocation and glucose uptake in skeletal muscles^3^, and enhanced metabolic flexibility reflected by increased capacity for fuel storage and substrate utilization^10^. Nonetheless, molecular mechanisms underlying the systemic health benefits of PA are not yet fully understood^11,12^.

Addressing this gap, plasma metabolomics in large cohorts enables the identification of metabolic signatures of PA and can highlight candidate biological pathways underlying these adaptations. While acute metabolic responses to exercise are typically examined under tightly controlled conditions and are well characterized for the blood metabolome^13–15^, the long-term metabolic adaptations to habitual PA have been studied less extensively, with studies typically relying on self-reported activity assessed using questionnaires, which yield less precise and potentially biased PA data^16^. In contrast, accelerometers provide more objective measures of movement intensity and duration but are less commonly applied in observational studies of free-living populations. The most consistent finding across these studies is that higher habitual PA is associated with lower circulating levels of branched-chain amino acids (BCAAs) and glutamate^17–25^.

The majority of metabolomics studies on habitual PA have been conducted in adults^17–21,26,27^, with limited data in children. In a Finnish cohort of girls, higher reported leisure-time PA at 11 years was associated with lower circulating isoleucine and leucine^24^. A similar inverse association with isoleucine was observed in another independent Finnish cohort of adolescents aged 13–19 years, where higher PA was also associated with lower lactate and higher levels of lipid-related metabolites, including phosphatidylcholines, sphingomyelins, and polyunsaturated fatty acids^23^. In a UK-based study, higher accelerometer-measured PA between 12 and 15 years was associated with lower pyruvate and alanine levels, and higher citrate and phenylalanine levels^25^.

In this study, we aimed to deeply characterize the link between habitual PA, sedentary behavior, and the plasma metabolome in children. For this purpose, we investigated the associations between plasma metabolite levels, quantified through targeted metabolomics, and five PA variables, assessed by accelerometry, in children aged 5.5-11 years from the European Childhood Obesity Project (CHOP) cohort. We validated our findings in the Toddler Milk Intervention trial (ToMI) cohort, with both cohorts comprising participants from different European countries. We further examined whether PA-metabolite associations vary according to age, sex and body mass index (BMI), three factors that themselves associate with PA and with metabolite profiles independently of PA.

## Methods

### Study design and population

The present study investigated cross-sectional associations between PA and the plasma metabolome at multiple childhood ages within CHOP^28,29^, a longitudinal cohort, and replicated the analyses in the independent longitudinal cohort of the ToMI trial^30^.

Both studies were originally initiated as randomized controlled trials to investigate the effects of different protein levels in infant formula during the first (CHOP) and second (ToMI) year of life on body composition later in life. After the intervention, the children of both studies underwent multiple follow-up assessments. Detailed descriptions of the two studies are provided in **Supplementary Material**.

In the present study, we used anthropometric, plasma metabolomic, and PA assessment data from 515 CHOP children, collected at 5.5 (or 6), 8, and 11 years of age. Each child contributed one to three complete observations. For our validation analysis in ToMI, we used anthropometric, plasma metabolomic, and PA data obtained for 683 children at 2 and/or 6 years. In both cohorts, information on participants’ sex and country of residence was available. BMI was calculated from weight and height measurements at 6, 8, and 11 years in CHOP and at 2 and 6 years in ToMI. To account for sex- and age-specific differences in body composition, zBMI scores were calculated using AnthroPlus software^31^.

All research was conducted in accordance with the Declaration of Helsinki and the standards of good clinical practice. Ethics committees at each study center approved the BiomarKid study protocol (IISPV 095/2022, LMU Ethik-Kommission-Nr: 02070 & Project Nr: 555-15 UMIL 0009763, CHC Liege 13/48/671, CHMI n°682). Written informed consent from parents or legal guardians was obtained for children in both studies. In CHOP, the children’s consent was additionally obtained at 8 and 11 years.

### PA assessment

Habitual PA in CHOP was assessed at 6, 8, and 11 years using the SenseWear Armband 2 (Body Media Inc., Pittsburgh, PA). At each age, the device was worn over the right triceps muscle for at least 20 hours per day on three consecutive days. To be included in the study, the participants needed measurements from at least one weekday and one weekend day. Movement was recorded in 1-minute epochs. Data preprocessing was performed with Professional InnerView Software 6.1^32^. PA measurements were categorized into four intensity levels using metabolic equivalents of task (METs). Light PA (1.5 to < 4 METs), moderate PA (4 to < 6 METs), and vigorous PA (6 to < 9 METs) were defined according to recommendations by Trost et al^33^, while very vigorous PA was defined as ≥ 9 METs. Additionally, total PA was defined as duration of all activities ≥ 1.5 METs and moderate-to-vigorous PA as duration in activities ≥ 4 METs. Sedentary behavior was defined as time spent in activities < 1.5 METs, after excluding periods recorded as lying and sleeping. Additional details and the rationale for the PA assessment steps have been published previously^34,35^. For this study, PA levels [min/day] were normalized for device-wear-time by dividing each child’s activity time by their device-wear-time and multiplying it with the mean device-wear-time across all children at all ages. Due to the presence of zero and identical values for vigorous and very vigorous PA across participants, these two PA parameters were categorized into tertiles and further analyzed as numerical variables (1, 2, 3).

In ToMI, PA was assessed at 2 and 6 years with the ActiGraph wGT3X-BT accelerometer (ActiGraph LLC, Pensacola FL, USA). The ActiGraph was used with idle sleep mode and a sampling rate of 30 Hz. It was worn on the right hip for 7 consecutive days and taken off only for sleeping or water-based activities. If the device was not worn for at least 4 days, with a minimum of 6 hours of wear time per day, then the individual was excluded. Data from the ActiGraph device were imported into R with the *read.gt3x* package^36^ and converted into activity counts (a measure of accelerometer-derived movement intensity) using the *agcounts* package^37^. Non-wear periods were identified and removed using the Choi algorithm^38^. Activity count data were categorized separately for measurements at 2 and 6 years of age, using the cut-points suggested by Trost et al. (sedentary behavior: ≤ 25 ActiGraph acceleration intensity counts; total PA: > 25 counts; moderate-to-vigorous PA: ≥ 420 counts; epoch = 15s)^39^ and Evenson et al. (sedentary: ≤ 25 counts; light PA: 26–573 counts; moderate PA: 574–1002 counts; vigorous PA: ≥ 1003 counts, moderate-to-vigorous PA: ≥ 574 counts, epoch = 15s)^40^, respectively. Lastly, data was normalized for possible differences in device-wear-time, by dividing each child’s duration of PA by their wear-time and multiplying it with average wear-time of all children. Further details on PA data preprocessing in ToMI were described previously^41^. For the present study, we used only sedentary behavior, total PA, and moderate-to-vigorous PA from ToMI, to ensure comparability between ages.

### Blood sample collection

At each study site, blood samples for metabolite profiling were collected in EDTA tubes at 5.5, 8, and 11 years in CHOP and at 2 and 6 years in ToMI. After centrifugation (1500g for 10 min, at 4 – 8 °C), plasma samples were stored at -80 °C.

Fasting status was recorded at blood collection. In CHOP, samples were collected in the fasted state (> 6 hours after the last meal), with the exception of samples from German participants at ages 5.5 and 8 years. In ToMI, the time since last meal was recorded at both time points and categorized as: < 3 hours, 3–6 hours, or > 6 hours. At age 2 years, most samples were obtained after fasting for either 3–6 hours (45%) or more than 6 hours (45%), whereas at age 6 years, most samples were collected after > 6 hours of fasting (74%). Besides fasting status, season at blood collection (summer, autumn, winter, spring) was recorded.

### Metabolite profiling

Metabolite profiling of plasma samples (CHOP and ToMI, from all study centers) using tandem mass spectrometry coupled with liquid chromatography (LC-MS/MS) was conducted at the MS-Core Facility of the LMU Hospital Munich (Dr. von Hauner Children’s Hospital). Following the longitudinal study design, CHOP samples were measured after completion of each study visit. For ToMI, 2- and 6-year samples were collected and analyzed together in one sequence. For both, CHOP and ToMI, quality control (QC) samples were co-analyzed in six wells of each 96-well plate batch. For CHOP, adult human plasma was used as QC, whereas for ToMI, a pooled sample comprising plasma from 2- and 6-year samples was used as QC. For both studies, two commercial adult human control plasma samples (CP-I and CP-II; Recipe) were also co-analyzed in duplicate on each analytical well plate. Below, we briefly summarize the applied analysis steps, which were similar for the two cohorts. Detailed information on protocols is provided in the **Supplementary Material**. An extended description of calibration standards, internal standards, MS-parameters, Q1/Q3 transition, chromatographic parameters and quantitation can be found in the Supporting Information of Hao et al.^42^.

#### Sample preparation

For metabolite extraction, 50 µL of thawed plasma samples were added to 450 µL of internal standard mixture in methanol. The 96-well plates with the clear metabolic extracts were stored at -20 °C (CHOP) or -30 °C (ToMI) before LC-MS/MS.

#### HPLC-ESI-QTRAP-MS/MS

Different analytical methods were used for (i) targeted amino acid analysis after derivatization with n-butanol, (ii) targeted organic acid analysis, including α-aminoadipic acid and taurine, (iii) flow-injection-analysis (FIA) of phospholipids (phosphatidylcholines (PCs), lysophosphatidylcholines (LPCs), sphingomyelins (SMs)), in ToMI and CHOP, and acylcarnitines and hexoses in CHOP samples, (iv) targeted acylcarnitine analysis of ToMI samples after derivatization with n-butanol, (v) targeted homocysteine and cysteine analysis in CHOP, and (vi) free fatty acids (FFAs) analysis in CHOP.

#### Data acquisition and data processing

For CHOP samples, Analyst 1.5.1 (Sciex) was used for data acquisition as well as quantitation. For ToMI samples, Analyst 1.6.3 (Sciex) was used for acquisition and MultiQuant 3.0 (Sciex) and Excel (Microsoft Office) were used for quantitation. Peak integration, background subtraction, isotope correction, and quantitation of FIA-MS/MS results were performed with an in-house R-script using R statistical software (version 4.1.0). For lipids, in particular, PCs, LPCs, and SMs, only the neutral loss corresponding to the head group and the total mass-to-charge ratio (*m/z*) of the molecule were determined, i.e., the length, desaturation, site and bond type of the linked fatty acid residues were not directly characterized. The ‘a’ and ‘e’ in the labels of these lipids describe acyl and ether bonds, respectively, and were chosen under the assumption that even-chain fatty acids and lower degrees of desaturation are more common than odd-chain fatty acids or highly unsaturated species. The x:y after the lipid abbreviations denotes the total number of carbons (x) and double bonds (y) of all fatty acid chains in the lipid. Consequently, a variety of lipid species with the same *m/z* and neutral loss are measured under any specific label as has been shown previously^43^. As an example, the analyte labeled PC ae C40:1 is a sum measure of all isobaric PCs, which, among other PCs, include acyl-alkyl (ae) PCs with 41 carbons and 8 double bonds (41:8), diacyl PCs (aa) with 39 carbons and 1 double bond (39:1) and diacyl PCs (aa) with 40 carbons and 8 double bonds (40:8) when considering a mass resolution of < 0.7 Da). Metabolites that could not be confidently annotated were labeled as unknown, followed by the precursor ion *m/z* (e.g., Unknown_458.3).

### Metabolomics data preprocessing and harmonization

A multi-step QC procedure has been applied to metabolomics data from CHOP and ToMI separately. Detailed descriptions of all steps are provided in **Supplementary Material**. Briefly, we checked the data set for sample outliers, performed batch correction using QC samples and estimated the remaining technical variation. Metabolites were excluded if they met the following criteria: (i) a coefficient of variation > 30% in CP-II samples (or in QC samples for acylcarnitines and phospholipids), (ii) a dispersion ratio, defined as the standard deviation of QC samples divided by that of study samples, greater than 50%. Metabolites with missingness exceeding 30% were excluded for the present analysis. Finally, missing values were imputed using a random forest approach.

In CHOP, the final data set included 24 amino acids (including homocysteine and cysteine (on 2 different platforms)), 18 organic acids, 50 free fatty acids (FFAs), hexoses, 26 acylcarnitines (including free carnitine), 28 LPCs, 106 PCs, 37 SMs, as well as 25 unidentified lipid metabolites labelled in this study by their *m/z*.

In ToMI, the final data set included 22 amino acids, 15 organic acids, 60 acylcarnitines (including free carnitine), and 167 phospholipids (91 PCs, 25 LPCs, and 51 SMs).

### Statistical analysis

Before statistical analysis, metabolite outlier values were winsorized using a threshold of five times the interquartile range (Q1 - 5×IQR and Q3 + 5×IQR). On the winsorized data, 49 biologically relevant metabolite sums and 42 metabolite ratios were calculated. Lastly, all metabolite values were log2-transformed and scaled separately for each age group and cohort. No outliers were identified for any PA variables based on the 5×IQR criterion. Correlations between PA variables in CHOP were assessed using Spearman correlation. Numerical PA variables and zBMI were scaled before statistical modelling.

All statistical analyses were performed using R (version 4.4.0).

#### PA association analyses

We examined the association between metabolite levels and PA variables in the CHOP cohort by fitting linear mixed-effects models using the lmerTest R package (version 3.1-3) with restricted maximum likelihood estimation (REML = TRUE) and Satterthwaite approximation for degrees of freedom. To assess the assumption of linearity between metabolite levels and PA, we compared linear mixed-effects models with and without an additional quadratic term for continuous PA variables. Models were compared using likelihood ratio tests and differences in Akaike information criterion (ΔAIC), with false discovery rate (FDR) correction applied to account for multiple testing. Linearity of associations was assessed and confirmed for all models.

In CHOP, the analysis was performed for 315 individual metabolites, 49 metabolite sums, and 42 metabolite ratios as outcomes (**Supplementary Table 1**). Each metabolite was modelled against five PA measures: sedentary behavior, total, moderate, vigorous, and very vigorous PA. Light PA was not included in the main analysis as we considered total PA as a proxy for predominantly low-intensity PA (**Supplementary Fig. 1A**). Sex, zBMI, and season were included as fixed effects, and visit, subject, and country as random effects. Country was included as a random effect, as it has previously been shown to explain substantial metabolomic variation in CHOP^44^. For 41 metabolites with reliable measurements (i.e., passing QC) at only one visit, subject and visit random effects were omitted from the models.

The FDR across 2,030 tests in CHOP (5 PA variables × 406 metabolite measures) was controlled using the Benjamini–Hochberg procedure, and FDR-adjusted p-values (q-values) < 0.05 were considered statistically significant. Metabolite associations were considered informative if (nominal) p values were < 0.05. For metabolite sums and ratios, an association was considered informative if p < 0.05 and the association of the ratio or sum had a lower p-value than the associations for their constituent metabolites (p-gain = min(p of constituents)/p of ratio or sum > 1)^45^.

#### Sensitivity analyses

For sensitivity analysis, information on puberty status (prepubertal and pubertal) was added as an additional fixed effect in the linear mixed-effects models in CHOP. Pubertal status was assessed at age 11 years using the Pubertal Development Scale^46^. Children at 5.5 and 8 years were considered prepubertal, whereas at 11 years, 97 children were recorded as prepubertal and 258 as pubertal (including early, mid, and late pubertal). Subjects with no puberty information at 11 years (n = 6) were excluded from this sensitivity analysis.

As an additional sensitivity analysis, fish intake was added as a fixed effect in models for metabolites presumably containing docosahexaenoic acid (DHA, 22:6), which is highly abundant in fish. These metabolites were LPC a C22:6, PC aa C38:6, PC aa C40:6, and PC aa C42:6^43^. Fish intake was assessed at ages 5.5 and 8 years using 3-day dietary records and at age 11 years using a food frequency questionnaire. Analysis was restricted to the subset of samples with dietary data (n = 775), and models without fish adjustment were refitted in the same subset for comparison.

#### Replication analysis in ToMI

For validation of our findings in the independent ToMI cohort, we performed the association analysis for all metabolites that were associated in CHOP with the corresponding PA variable (for moderate-to-vigorous PA: with moderate, vigorous or very vigorous PA) and that were also reliably measured in ToMI. Metabolites were modelled against three PA measures (sedentary behavior, total, and moderate-to-vigorous PA) using the same linear mixed-effects models as in CHOP with the exception that fasting duration (< 3 h, 3-6 h, > 6 h) was included as an additional fixed effect. The levels of the three PA measures did not differ significantly between the two ToMI intervention groups (all p > 0.05). In total, 67 association tests were conducted (11 for sedentary behavior, 16 for total PA, 40 for moderate-to-vigorous PA). Associations that were nominally significant and showed concordant directions of effect in CHOP and ToMI were considered replicated. P-values were adjusted (q values) for multiple testing across 67 tests using the Benjamini–Hochberg procedure. To assess overall concordance between PA effect estimates in ToMI and CHOP cohorts, we additionally applied linear mixed-effects models to model the association of all reliably measured metabolites available in both cohorts (n = 177) with sedentary behavior, total PA, and moderate-to-vigorous PA, respectively. Metabolite sums and ratios were not included in this comparison. The correlation between β effect estimates derived from the ToMI and CHOP cohorts was quantified using Pearson’s correlation coefficient (r). Statistical significance of the correlation was evaluated using stats::cor.test (R version 4.4.0), with p-values calculated based on the Student’s t-distribution.

#### Interaction and stratified analyses

To assess whether associations between PA and metabolite levels differed by sex and BMI, we fitted interaction models including *PA* × *sex* and *PA* × *zBMI* terms in CHOP. For BMI interaction analysis, significant (p < 0.05) interactions were visualized using the interactions::interact_plot (version 1.2.0) function in R, displaying predicted metabolite levels across tertiles of the moderator (modx.values = “terciles”) with 80% confidence bands (int.width = 0.80).

Additionally, linear mixed-effects model analyses were performed stratified by sex and by age/visit. For sex-stratified analysis, sex was removed as fixed effect, while for visit-stratified analysis, visit and subject were removed as random effects. Sex-stratified results were visualized as dot plots displaying metabolite-specific effect estimates separately for males and females, generated using the *ggplot2* (version 4.0.2) and *patchwork* (version 1.2.0) packages in R. Metabolites reliably measured only at one visit and metabolite sums and ratios were not included in the plot. For age-stratified analyses, only (single) metabolites measured at all three time points were included. To account for unequal sample sizes across visits, analyses were standardized to 280 samples, corresponding to the sample size of the 5.5-year visit. For the 5.5-year visit, the model was run once. For the 8- and 11-year visits, which had larger sample sizes, we conducted 100 down-sampling iterations, each time randomly selecting 280 samples. Mean and SD of the number of significantly associated metabolites (p < 0.05) across 100 iterations and per visit were calculated. The difference in the number of significant hits at 8 and 11 years was tested with a paired Wilcoxon signed-rank test across 100 matched resampling iterations.

To identify sex- and BMI-associated metabolites, we fitted linear mixed-effects models without PA variables using sex and zBMI as fixed effects and otherwise including the same covariates as in the PA association models.

#### Metabolite set enrichment analysis

To obtain an overview of metabolite associations with PA on the level of metabolite classes, we performed a metabolite set enrichment analysis for association tests performed above, i.e., β estimates for PA from each of the main models, β estimates for PA × sex from each interaction model, and β estimates for sex or zBMI from the corresponding models without PA. These β estimates were used to rank metabolites for the enrichment analysis. Metabolite sets were defined based on metabolite classes (see **Supplementary Table 1**). Metabolites with reliable measurements at only one visit were excluded from the analysis, as β estimates from such models were not comparable to those obtained from models including visit and subject as random effects. The analysis was performed using fgsea::fgseaMultilevel function (version 1.30.0), with parameters set to minSize = 2, maxSize = 500, and nPermSimple = 10000.

## Results

In this study, we examined the relationship between plasma metabolite levels and habitual PA in 515 children of the CHOP cohort recruited from five European countries. Specifically, we investigated metabolite associations with five accelerometry-assessed PA variables: sedentary behavior, total PA, moderate PA, vigorous PA, and very vigorous PA. Light PA, which accounted for approximately 80% of total PA on average, was not included as a separate PA variable in the main analysis; instead, we considered total PA as a proxy (results for light PA are provided in **Supplementary Table 2**). The five PA variables were examined in relation to 406 metabolite variables, including 315 individual metabolites quantified by targeted metabolomics, 42 ratios, and 49 sums. Identified associations were subsequently tested for replication in the independent child cohort ToMI. Finally, we studied whether metabolite associations interacted with sex or BMI and whether they varied across ages.

### Population characteristics

In CHOP, metabolite profiles were assessed at 5.5, 8, and 11 years, with 120 children being profiled at all three visits, 203 at two, and 192 at one. Participant characteristics are presented in **Table 1**. With age, sedentary time increased (Spearman ρ = 0.45, p < 2.2 × 10⁻^16^) and total PA decreased (ρ = -0.33, p < 2.2 × 10⁻^16^). Engagement in moderate, vigorous, and very vigorous activity was highest at 8 years. Sedentary time was negatively correlated with all other PA measures (ρ = -0.26 (very vigorous PA) to -0.84 (total PA)), while moderate, vigorous, and very vigorous PA measures were positively correlated with each other (ρ = 0.40 to 0.63) (**Supplementary Fig. 1B**).

**Table 1.**
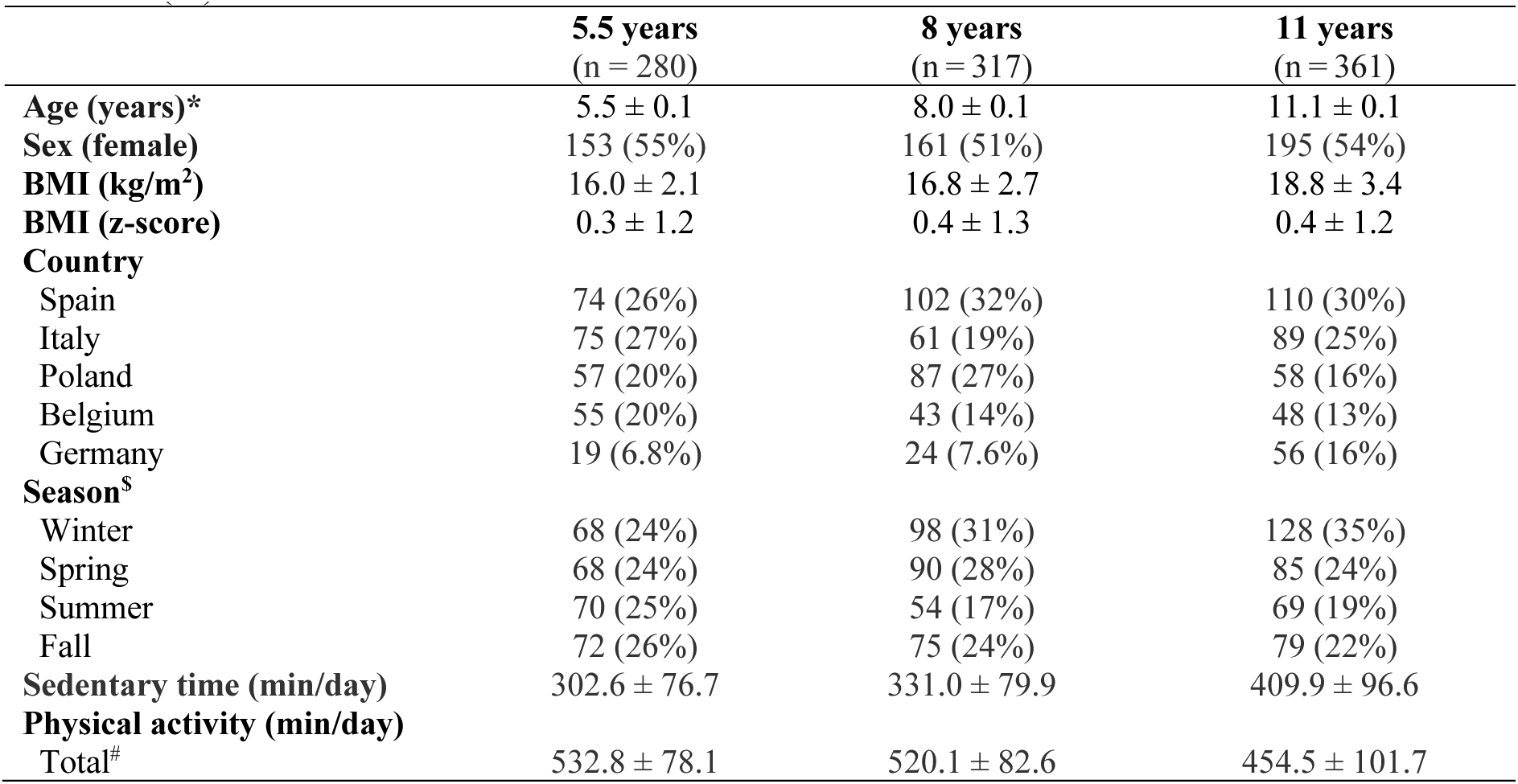
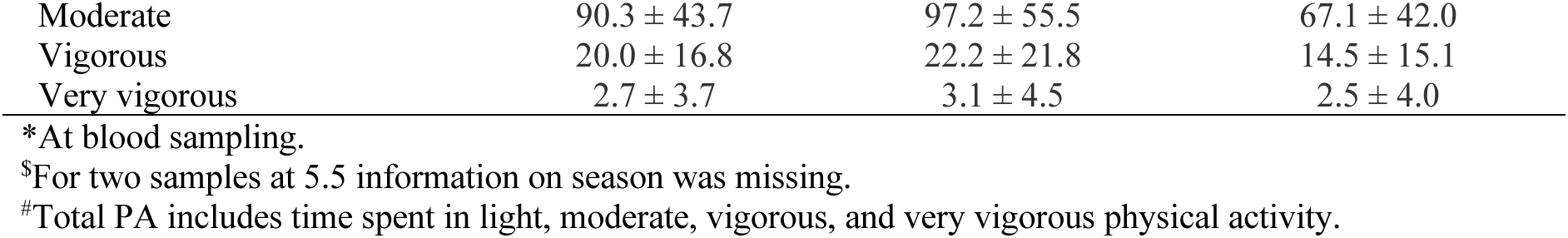
Characteristics of the CHOP cohort used in this study. Values are presented as mean ± SD or n (%).

### Associations between metabolites and habitual physical activity

To assess the relationship between metabolite levels and habitual PA, we applied linear mixed-effects models separately for each of the five PA measures (adjusted for sex, zBMI and season as fixed effects, and including subject, visit, and country as random effects). Of the 315 single metabolites tested, 62 were associated with at least one PA measure (p < 0.05). In addition, three of 49 metabolite sums and eight of 42 metabolite ratios met the significance criteria (p < 0.05 and p-gain > 1), resulting in a total of 73 metabolite variables associated with PA (**Fig. 1, Supplementary Table 2)**. After correction for multiple testing, none of the associations remained significant at a false discovery rate (FDR) of q < 0.05. Results were very similar when we added puberty status as an additional covariate (**Supplementary Table 2)**.

**Fig. 1.**
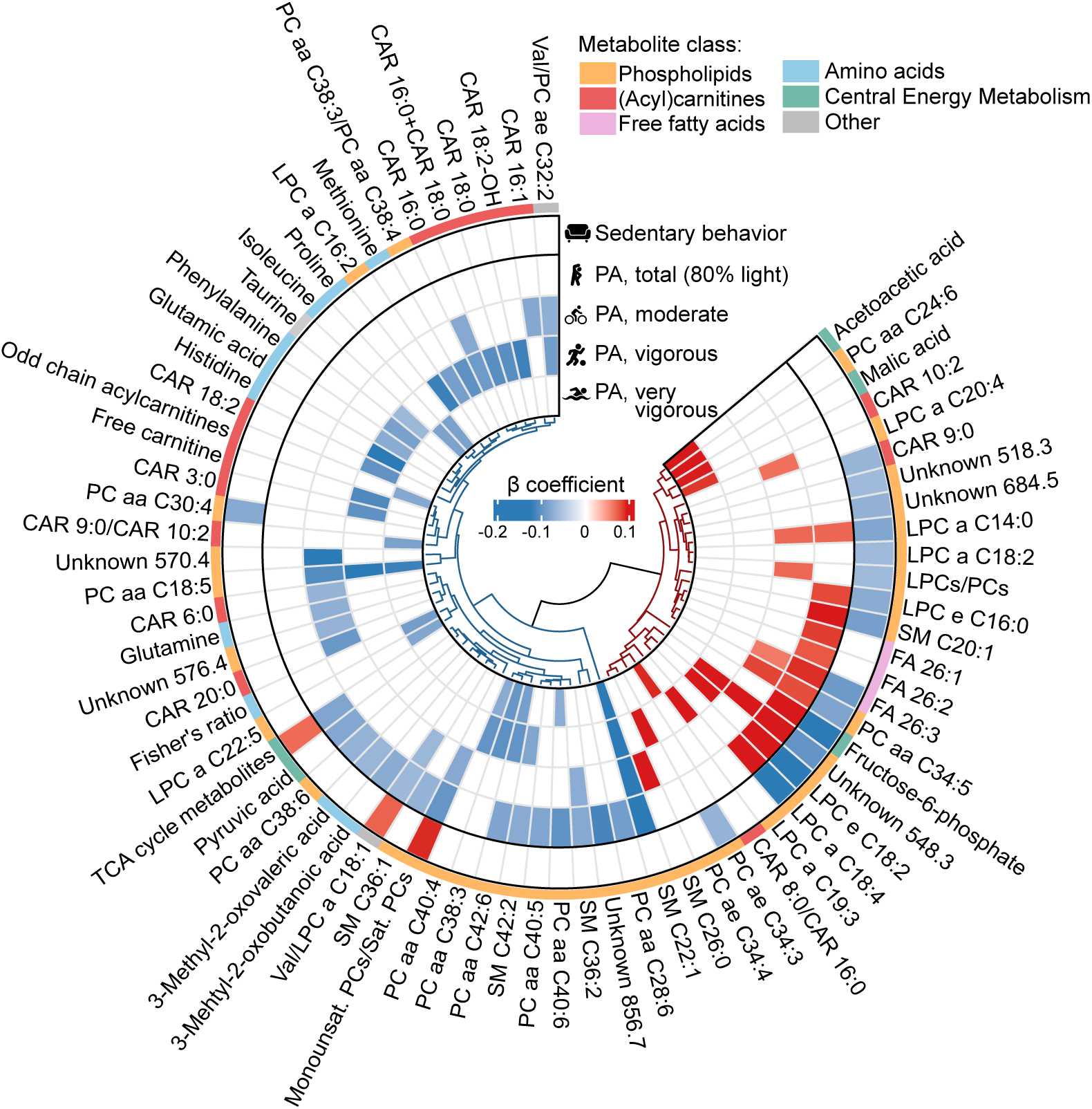
Associations between plasma metabolite levels and physical activity (PA) measures. The circular heatmap displays individual metabolites as well as metabolite sums and ratios that were significantly (nominal) associated with at least one of the five PA measures based on linear mixed-effects models including all ages (5.5, 8, 11 years). Nominal significance was defined as p < 0.05 for individual metabolites and as p < 0.05 with p-gain > 1 for metabolite sums and ratios. Grid colors represent β estimates from the linear mixed-effects models, with the color scale ranging from the minimum to the maximum β values across all models and metabolite variables available at all three time points. Non-significant associations are shown in white. Metabolites are ordered by hierarchical clustering, based on all β estimates, including non-significant ones. The outer color band indicates metabolite class. Monounsat.: monounsaturated, Sat.: saturated.

Of the 73 associated metabolite variables, 46 (63%) were lower in children with more PA (**Fig. 1**). For metabolites associated with multiple PA measures, the direction of effect was consistent across total PA and the three PA intensities and opposite to that observed for sedentary behavior. The number of associations (p < 0.05) was highest for total PA (n = 27), followed by vigorous PA (n = 25), moderate PA (n = 20), sedentary behavior (n = 19), and very vigorous PA (n = 17). Among PA variables, the largest overlap in associated metabolites was observed for sedentary behavior and total PA (n = 12). In contrast, very small (n = 2) or no overlap was observed between sedentary behavior and vigorous or very vigorous PA, respectively.

Metabolite set enrichment analysis of metabolite ∼ PA associations (**Supplementary Table 3**) showed enrichment of negative associations in amino acids with higher PA intensities (q = 0.032 – 0.037). A similar pattern was observed for acylcarnitines. Most associated acylcarnitines were lower in more active children, with significant enrichment found for moderate (q = 7.9 x 10^-7^) and vigorous PA (q = 0.032). Negative (positive) associations with total PA (sedentary behavior) were enriched for the group of BCAAs and their derivatives (q = 0.047 and q = 0.029, respectively) though only the two alpha-ketoacids 3-methyl-2-oxobutanoic acid and 3-methyl-2-oxovaleric acid were nominally significantly associated. Out of 50 measured FFAs, only FA 26:3 was associated with total and moderate PA, and (inversely) with sedentary behavior, and FA 26:1 and FA 26:2 with total PA. In the enrichment analysis, however, FFAs were enriched for positive associations with very vigorous PA. LPCs were enriched for higher levels with moderate and total PA and lower levels with sedentary behavior (q = 3.4 x 10^-4^ – 0.047). Four LPCs (LPC a C14:0, LPC a C18:4, LPC e C16:0, LPC e C18:2) showed both significant (nominal) positive associations with total PA and negative associations with sedentary behavior. For PCs, negative associations with sedentary behavior were enriched (q = 0.039) but no coherent direction of associations was seen with the remaining PA variables. Notably, several DHA-containing PCs, such as PC aa C42:6, PC aa C40:6, and PC aa C38:6, were consistently lower in children with higher total PA. Effect estimates remained similar after additional adjustment for fish intake (**Supplementary Table 4**). Among the metabolites of the central energy metabolism, levels of malic acid and acetoacetic acid were higher in children with more very vigorous PA, and pyruvic acid was lower in those with more total PA. No significant enrichment was found for this group.

In addition to single metabolites, several ratios previously associated with higher BMI or insulin resistance^47,48^ were nominally significantly lower in more physically active children, including Fisher’s ratio (BCAAs/aromatic amino acids)^49^, and two ratios with valine (Val), namely Val/LPC a C18:1, and Val/PC ae C32:2.

### Replication of associations in ToMI

To validate our findings, we replicated the analyses in the ToMI cohort, in which associations between plasma metabolites and accelerometer-measured habitual PA were assessed in 683 children at ages 2 and/or 6 years (895 observations). Characteristics of the ToMI population included in these analyses are provided in **Supplementary Table 5**. Validation analyses included three PA variables: sedentary behavior, total PA, and moderate-to-vigorous PA, with the latter analyzed as a combined measure because moderate, vigorous, and very vigorous PA were only jointly assessed at age 2 years in ToMI. We performed replication analysis for all metabolites that were associated with PA in CHOP (p < 0.05) and reliably measured in the ToMI cohort (**Supplementary Table 6**).

Of the eleven metabolites available for replication analysis for sedentary behavior, five replicated in ToMI (**Table 2**). For total PA, six of the 16 tested metabolite variables were replicated. For both activity measures, replicated metabolites showed concordant effect directions across cohorts. For the metabolite variables associated with either moderate, vigorous or very vigorous PA in CHOP, 40 were tested against moderate-to-vigorous PA in ToMI, of which nine reached significance (p < 0.05, p-gain > 1). However, two metabolites (histidine and glutamine) showed discordant directions of association, therefore, six metabolites were considered successfully replicated. The association of the monounsaturated PCs/saturated PCs ratio was replicated across all three PA variables. In addition, LPC a C14:0, PC aa C40:6, LPCs/PCs, and Val/LPC a C18:1 were associated with more than one PA variable in both cohorts. The strongest replicated associations were observed for LPCs/PCs with moderate-to-vigorous PA (q = 8.1×10⁻^4^), PC ae C34:3 (q = 2.8 ×10⁻^3^) and CAR 9:0 (q = 4 .2 ×10⁻^2^) with sedentary behavior, and LPC a C14:0 with total PA (q = 4.2 ×10⁻^2^).

**Table 2.** Validation of CHOP-discovered metabolite associations with physical activity (PA) variables in the ToMI cohort.

| **Metabolite** | **ToMI<br>(2, 6 years)** |  |  |  | **CHOP<br>(5.5, 8, 11 years)** |  |  |  |
|---|---|---|---|---|---|---|---|---|
|  | **β** | **p** | **p-gain** | **q** | **β** | **p** | **p-gain** | **q** |
| **Sedentary behavior** |  |  |  |  |  |  |  |  |
| **PC ae C34:3** | -0.1265 | 1.2705×10<sup>−4</sup> |  | 0.0028 | -0.0714 | 0.0434 |  | 0.8445 |
| **CAR 9:0** | -0.0962 | 0.0032 |  | 0.0424 | -0.0692 | 0.0321 |  | 0.8336 |
| **LPCs/PCs** | -0.0872 | 0.0120 | 2.0167 | 0.0940 | -0.0651 | 0.0443 | 1.2733 | 0.8445 |
| **Monounsat. PCs/Sat. PCs** | 0.0730 | 0.0225 | 16.7119 | 0.0940 | 0.0901 | 0.0139 | 57.9396 | 0.6760 |
| **Val/LPC a C18:1** | 0.0716 | 0.0391 | 3.6555 | 0.1377 | 0.0708 | 0.0243 | 2.2038 | 0.7479 |
| **Total PA** |  |  |  |  |  |  |  |  |
| **LPC a C14:0** | 0.0964 | 0.0030 |  | 0.0424 | 0.0693 | 0.0349 |  | 0.8336 |
| **Val/LPC a C18:1** | -0.0785 | 0.0189 | 3.6860 | 0.0940 | -0.0658 | 0.0362 | 3.4087 | 0.8345 |
| **SM C36:1** | -0.0852 | 0.0206 |  | 0.0940 | -0.0713 | 0.0396 |  | 0.8445 |
| **PC aa C40:6** | -0.0706 | 0.0283 |  | 0.1113 | -0.1035 | 0.0029 |  | 0.6760 |
| **LPC e C16:0** | 0.0761 | 0.0345 |  | 0.1285 | 0.0805 | 0.0113 |  | 0.6760 |
| **Monounsat. PCs/Sat. PCs** | -0.0737 | 0.0448 | 3.4564 | 0.1500 | -0.0997 | 0.0040 | 49.4018 | 0.6760 |
| **Moderate-to-vigorous PA<sup>\*</sup>** |  |  |  |  |  |  |  |  |
| **LPCs/PCs** | 0.1476 | 1.2081×10<sup>−5</sup> | 19.7458 | 0.0008 | 0.0551 | 0.0960 | 1.1925 | <sup>#</sup> |
| Histidine<sup>\$</sup> | 0.1405 | 3.3387×10<sup>−5</sup> |  | 0.0011 | -0.0393 | 0.2629 |  | <sup>#</sup> |
| **PC ae C34:4** | 0.0872 | 0.0115 |  | 0.0940 | 0.0793 | 0.0221 |  | <sup>#</sup> |
| **CAR 10:2** | 0.0862 | 0.0132 |  | 0.0940 | -0.0179 | 0.6046 |  | <sup>#</sup> |
| **LPC a C14:0** | 0.0811 | 0.0147 |  | 0.0940 | 0.0675 | 0.0449 |  | <sup>#</sup> |
| **PC aa C40:6** | -0.0812 | 0.0159 |  | 0.0940 | -0.0473 | 0.1562 |  | <sup>#</sup> |
| **LPC a C20:4** | 0.0787 | 0.0177 |  | 0.0940 | 0.0550 | 0.1051 |  | <sup>#</sup> |
| **Monounsat. PCs/Sat. PCs** | -0.0770 | 0.0200 | 4.5255 | 0.0940 | -0.0677 | 0.0505 | 12.9668 | <sup>#</sup> |
| Glutamine<sup>\$</sup> | 0.0768 | 0.0224 |  | 0.0940 | -0.0722 | 0.0233 |  | <sup>#</sup> |

<sup>\*</sup>For moderate-to-vigorous PA, CHOP columns show estimates from the moderate-to-vigorous PA model, which was not part of the primary analysis. Metabolites were selected and directional concordance was assessed based on their nominally significant association with moderate, vigorous, or very vigorous PA in CHOP.
<sup>\$</sup>Metabolites associated with PA in both cohorts, but with opposite β coefficient direction.
<sup>#</sup>Non-adjusted p-values are reported for moderate-to-vigorous PA, since this combined PA variable was not included in the primary CHOP analysis, for which multiple testing correction was performed. The Benjamini-Hochberg procedure was applied to calculate FDR-adjusted q-values. Monounsat.: monounsaturated, Sat.: saturated.

When considering all metabolites reliably measured in both CHOP and ToMI, their PA β coefficients were significantly correlated for all three PA measures (**Supplementary Fig. 2**). The strongest correlation between β estimates from the two cohorts was observed for total PA (r = 0.309, p = 2.9 ×10⁻⁵), followed by moderate-to-vigorous PA (r = 0.280, p = 1.6 ×10⁻⁴), and sedentary behavior (r = 0.261, p = 4.4 ×10⁻⁴).

### Sex differences in the association between metabolites and physical activity

Next, we studied the influence of sex on the associations between metabolites and PA, using linear mixed-effects models with PA × sex interaction (using zBMI as one of the covariates) (**Supplementary Table 7**). To facilitate interpretation of sex interaction effects, we additionally performed a metabolite set enrichment analysis using β estimates from the sex × PA interaction **(Fig. 2A, Supplementary Table 8**) and sex-stratified analyses (**Fig. 2B, Supplementary Fig. 3, Supplementary Table 7)**. For further context, we also performed metabolite-sex association analyses in CHOP (**Supplementary Fig. 4 and Supplementary Tables 9** and **10**).

**Fig. 2.**
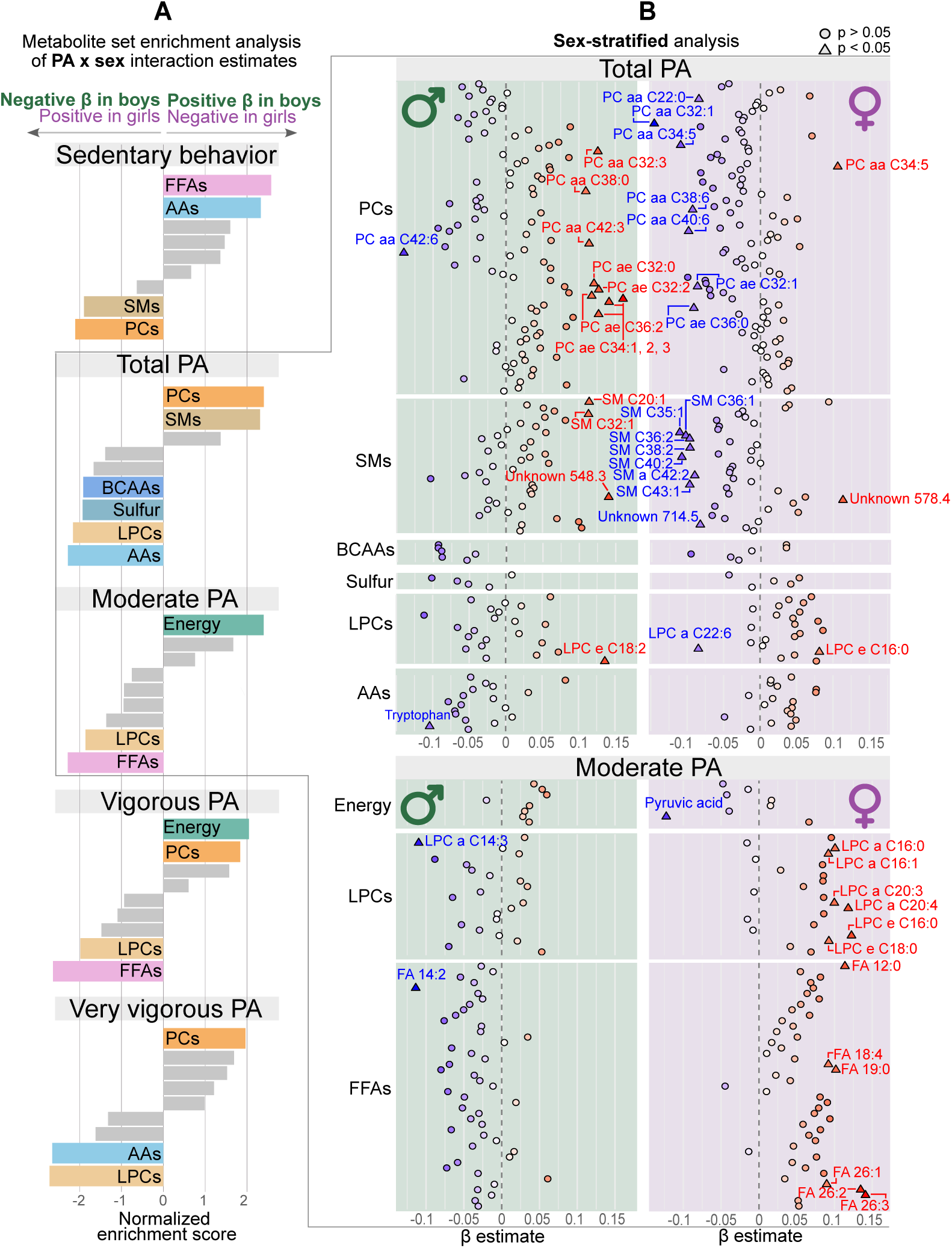
Influence of sex on associations between metabolite levels and physical activity (PA). **A)** Metabolite set enrichment analysis using β coefficients from the sex × PA interaction term in the linear mixed-effects models. Bars represent normalized enrichment scores for each metabolite group within each PA variable. Colored bars indicate metabolite groups with significant (q < 0.05) sex interactions, while grey bars indicate non-significant metabolite groups. **B)** Sex-stratified analysis of associations between metabolites and total or moderate PA. Color represents β estimates, with darker blue reflecting stronger negative and darker red stronger positive associations. Nominally significant associations (p < 0.05) are depicted in triangles and labeled with name, while not significant metabolites are depicted as circles. Metabolites reliably measured only at one visit and metabolite ratios and sums are not included in the plot. AA: amino acids; BCAA: branched-chain amino acids; Energy: metabolites involved in central energy metabolism (e.g., TCA cycle metabolites, glucose); FFAs: free fatty acids; LPCs: lysophosphatidylcholines; PCs: phosphatidylcholines; SMs: sphingomyelins; Sulfur: metabolites involved in sulfur metabolism (e.g., cysteine, homocysteine and taurine).

At nominal significance (p < 0.05, p-gain > 1), 45 metabolite variables showed sex interactions with sedentary behavior, 23 with total PA, 18 with moderate PA, 31 with vigorous PA, and 33 with very vigorous PA (**Supplementary Table 7**). The strongest result was the positive interaction of sedentary time and sex for Val/PC ae C32:2 (p = 8.20 ×10⁻⁵, q < 0.1) with higher values of Val/PC ae C32:2 being associated with more sedentary time in boys, while no significant effect was observed in girls in the stratified analysis.

In the enrichment analysis **(Fig. 2A)**, LPCs showed a significant enrichment of sex interactions across all PA measures except sedentary behavior. The strongest enrichment was observed for very vigorous PA (q = 1.98 ×10⁻⁵), for which the majority of LPCs were lower in boys with more very vigorous activity, whereas in girls no universal pattern was observed for LPCs. PCs showed significant enrichment for all PA measures except moderate PA. Sex-stratified analyses suggested that these enrichments were primarily driven by specific PC subgroups such as PCs with acyl alkyl (ae) linkages, some of which were higher in more active boys or lower in more active girls **(Fig. 2B)**. Conversely, FFAs were higher in more active girls and lower in more active boys. Central energy metabolism was enriched for PA × sex interactions with moderate and vigorous PA (q = 0.023 and 0.013). With moderate and vigorous PA, pyruvic acid was lower in more active girls but not in boys. With very vigorous PA, methylmalonic acid and malic acid were higher in more active boys only. β-Hydroxybutyric acid showed positive interactions with total, moderate and very vigorous PA, however its sex-stratified associations were not significant in either sex.

### BMI modulates the association between metabolites and physical activity

To investigate BMI-dependent differences in metabolite associations with PA, we used linear mixed-effects models of metabolite levels with a PA × zBMI interaction term (and sex added as one of the covariates). Nominally significant (p < 0.05, p-gain > 1) interactions were identified for 42 metabolites with sedentary behavior, 25 with total PA, 21 with moderate PA, 18 with vigorous PA, and 13 with very vigorous PA **(Supplementary Table 11)**. β-Hydroxybutyric acid was the only metabolite showing a zBMI interaction for all PA variables, with a positive interaction effect for all variables except sedentary time. A positive interaction indicates that the PA–metabolite association becomes more positive with increasing zBMI. Depending on the magnitude of the interaction, this may reflect a stronger, weaker, or opposite-direction association at different levels of zBMI. To facilitate interpretation, we stratified zBMI per tertiles and plotted predicted PA-metabolite association for each tertile (**Fig. 3A**). For β-hydroxybutyric acid, the interaction results indicate that higher levels were observed with more activity in children with higher zBMI, whereas the opposite trend (i.e., lower levels with higher activity) was seen in leaner children **(Fig. 3A)**. For several metabolite variables, we observed nominally significant BMI interactions for multiple PA variables: Valine, leucine, and phenylalanine showed negative interaction effects with total and moderate PA, suggesting that lower levels of these amino acids associate with higher activity in children with higher zBMI, while this effect is weaker or even in opposite direction in leaner children **(Fig. 3A)**. Similarly, multiple carnitines, including free carnitine, CPT1 ratio (free carnitine/[CAR 16:0 + CAR 18:0]), and CAR 3:0, showed negative interaction effects with total and/or very vigorous PA in children with greatest zBMI whereas associations were weaker or positive for leaner children. Conversely, CAR 18:2 demonstrated the opposite pattern, being inversely associated with moderate and vigorous PA in leaner children but not in those with higher zBMI. Among phospholipids, SM C36:3 showed zBMI interactions for total, moderate, and very vigorous PA, exhibiting more positive associations with PA at higher zBMI.

**Fig. 3.**
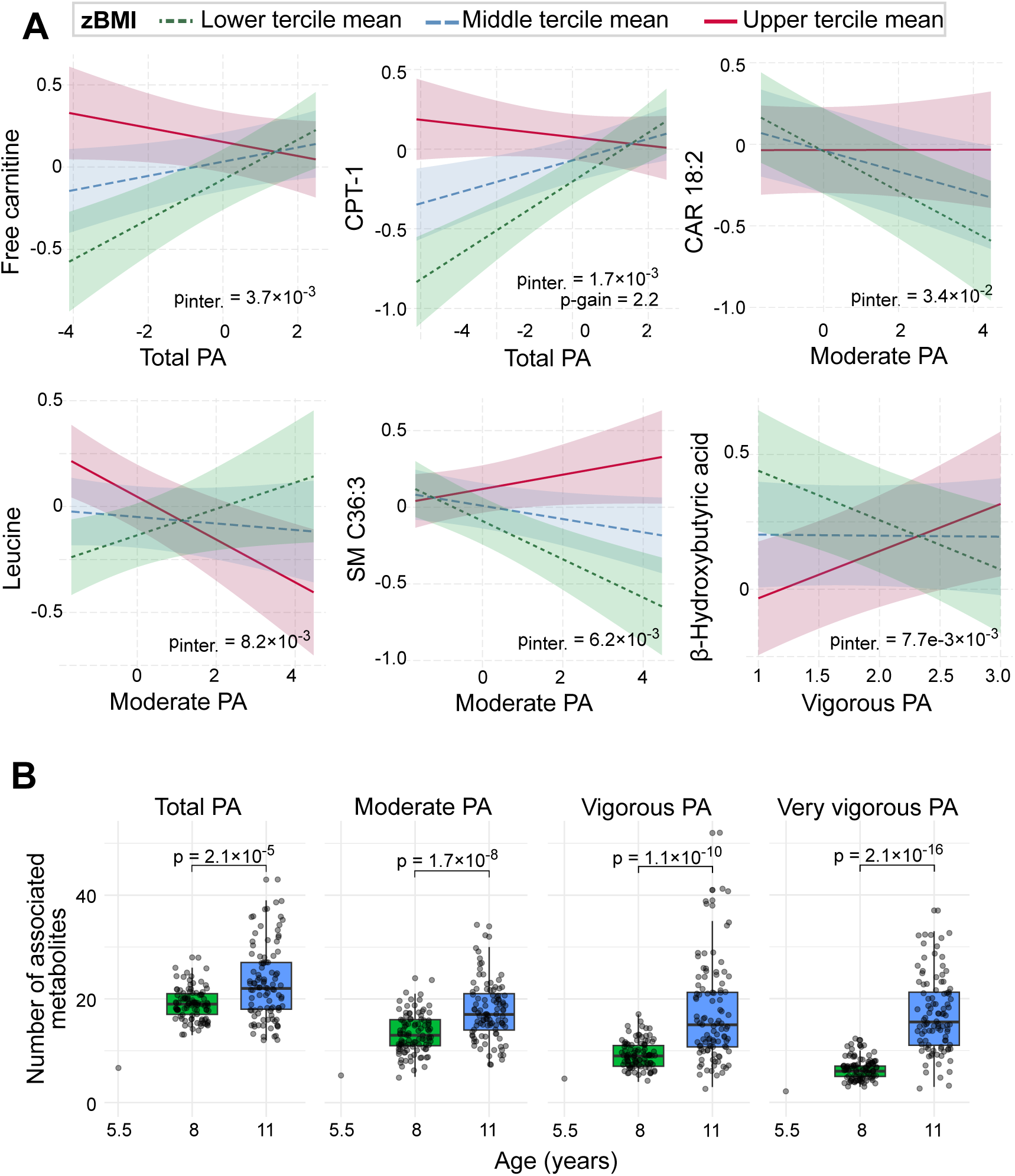
Impact of body mass index (BMI) and age on the associations between plasma metabolite levels and physical activity (PA). **A)** Interaction effect of zBMI on the associations between metabolite levels and PA. Interaction plots are based on linear mixed-effects models including a PA × zBMI interaction term. Lines represent predicted metabolite levels across different times spent in PA, stratified by zBMI tertiles, with sex and season held at reference levels and random effects (subject, visit, country) at population means (zero). Shaded areas indicate 80% confidence intervals. **B)** Number of PA-associated metabolites across the three ages. To ensure comparability across ages, samples at 8 and 11 years were randomly down-sampled to match the sample size at 5.5 years; this procedure was repeated 100 times. Linear mixed-effects models were fitted separately for each age and each down-sampling iteration. The figure shows the number of nominally significantly associated metabolites obtained in each of the 100 iterations for ages 8 and 11 years. At 5.5 years, no down-sampling was required, and the model was fitted only once. P-values are from paired Wilcoxon tests (8 vs 11 years), Bonferroni-corrected for five PA variables. CPT1: free carnitine/[CAR 16:0 + CAR 18:0].

Similar as for sex, we additionally assessed the associations between metabolites and zBMI for context: 84 metabolite variables (q < 0.05) were found associated with zBMI. Among others, DHA-containing lipids (e.g., PC aa C42:6, PC aa C38:6, and LPC a C22:6^43^), PC ae C34:3 and LPC a C18:2 were generally lower in children with higher zBMI. While Val/LPC a C18:1, CPT1 ratio and SM C34:2 were higher in those with higher zBMI (**Supplementary Fig. 5A, Supplementary Table 12**). In the metabolite set enrichment analysis, positive associations with zBMI were enriched for BCAAs and their derivates, and negative association for PCs **(Supplementary Fig. 5B, Supplementary Table 13)**.

### More associations with physical activity were observed in older children

To examine the consistency of PA associations across the three ages, we fitted age-stratified linear mixed-effects models and compared the number of nominally significant metabolite associations per age. To ensure that identified differences were not driven by the unequal sample sizes, we randomly down-sampled the datasets at ages 8 and 11 years to match the sample size at age 5.5 (n = 280). Across 100 down-sampling iterations, the mean number of associations increased with age for all PA intensities (**Fig. 3B, Supplementary Table 14**). In contrast, sedentary behavior showed the highest number of associated metabolites at age 5.5 and the lowest at age 8 (**Supplementary Fig. 6**).

## Discussion

In this metabolome-wide analysis, we show that device-measured habitual PA is associated with a metabolic imprint implicating various pathways, including lipid and BCAA metabolism, already in childhood. Associations for 12 metabolite variables replicated in an independent cohort, suggesting the consistency of these associations from toddlerhood to early puberty. The identified associations varied depending on PA intensity and became more numerous with age. Interestingly, PA associations partially differed between the two sexes and across the BMI range.

### Metabolite associations with habitual physical activity span various pathways

#### Carnitines were lower in children who exercised more

Children who spent more time in higher intensity activities, which include exercise, showed lower levels of free carnitine and multiple acylcarnitines. These metabolites are involved in transporting fatty acids from the cytosol into mitochondria for β-oxidation^50^. Their circulating levels are commonly used as markers of mitochondrial fatty acid oxidation, with higher levels often reflecting incomplete β-oxidation and lower levels suggesting greater metabolic efficiency^51–53^. Given that exercise is known to improve β-oxidation and mitochondrial capacity^9^, these associations may therefore reflect greater oxidative efficiency in children who exercise more.

#### Amino acids, including BCAAs and their degradation products, were lower in more active children

Similar to acylcarnitines, amino acids, including the BCAA isoleucine, were lower in children who spent more time in higher intensity activities. Moreover, two branched-chain α-keto acids, which are products of BCAA catabolism^54^, were inversely associated with total PA. In line with our results, multiple PA observational studies have reported lower BCAA levels in more active individuals in adulthood^18,21,22^ and childhood or adolescence^23–25^. Enhanced uptake and oxidation of BCAAs in contracting skeletal muscle during prolonged or extensive activity could explain these observations. Accordingly, activation of the branched-chain α-keto acid dehydrogenase complex, the second and rate-limiting step in BCAA oxidation, has been proposed as a potential mechanism^55^. Supporting this hypothesis, a twin study found that more active individuals exhibited higher expression of genes involved in BCAA degradation in both skeletal muscle and adipose tissue compared with their less active co-twins^56^. Also, the reason for lower plasma glutamate levels in individuals who exercise more, as observed in CHOP and seen consistently in adults^17–20^, could be connected to BCAA catabolism, as glutamate serves as a nitrogen acceptor in this process and in ammonia detoxification more generally^57^. In addition to higher BCAA oxidation, lower circulating BCAA levels in children who exercise more may reflect enhanced protein synthesis in muscle^58–61^.

#### LPCs, the products of PC hydrolysis, were higher in more active children

Various LPCs, including LPC a C14:0, were robustly associated with total PA and (inversely) with sedentary behavior. LPCs are generated from PCs - the most abundant phospholipids in mammalian cell membranes and lipoproteins - either through lecithin–cholesterol acyltransferase (LCAT), which transfers a fatty acid from PC to free cholesterol, producing cholesterol esters and LPC, or via hydrolysis of PC primarily by phospholipase A2 (PLA2), which removes one fatty acyl chain and releases an LPC^62^. Muscular PLA2 activity has been shown to increase after exercise in rats^63^, supporting a role for exercise-induced PLA2 activity. Consistent with this, we observed lower LPCs/PCs ratios in more sedentary children. In contrast to LPCs, we did not see a consistent pattern for PCs. We therefore hypothesize that in sedentary children, lipolytic activity is potentially reduced due to lower demand for lipid oxidation, potentially resulting in decreased PC hydrolysis and consequently lower circulating LPC levels and a reduced LPCs/PCs ratio.

Our findings are consistent with prior literature in adults. In intervention studies, LPCs, including LPC a C14:0, have been reported to increase with the exercise intervention^20,64^. Furthermore, in observational studies, higher plasma LPC levels have been reported in adults with more habitual PA after BMI adjustment^19^. Higher plasma LPC levels have also been associated with better mitochondrial function in skeletal muscle cross-sectionally^65^, and faster decline in plasma LPCs has been associated with faster decline in skeletal muscle mitochondrial function^66^. While it is important to keep in mind that the relationship between mitochondrial function and LPC levels is complex and not fully understood yet^67^, we speculate that the observed PA-related increases in circulating LPCs may contribute to the maintenance of mitochondrial function in active children.

#### DHA-containing PCs were lower in more active children

Some of the most consistent PA associations involved DHA-containing PCs, which were lower in more active children independent of fish intake, with PC aa C40:6 also replicating in ToMI. The same direction has been reported in adults, where lower levels of the corresponding phosphatidylethanolamine PE(40:6) were found in individuals with higher habitual PA^19^. Moreover, PC aa C40:6 levels decreased in a 10-week exercise intervention^20^. These observations may reflect a redistribution rather than a depletion of DHA in more active individuals. While greater weekly running distance in non-elite adults has been associated with a gradual decrease in the omega-3 index in blood^68^, regular exercise training has been shown to increase DHA content in phospholipids of skeletal muscle membranes^69^. Omega-3 fatty acids, including DHA, exert beneficial effects on skeletal muscle primarily through incorporation into membrane phospholipids, where they are associated with enhanced muscle protein synthesis and mitochondrial function^70,71^. A constant remodeling of fatty acid contents within PCs in relation to catabolic processes in skeletal muscle induced by fasting (transiently) or insulin deficiency has been shown in a mouse study. While these processes reduced DHA-containing PCs, the overall PUFA-containing PC pool remained stable^72^. This supports the hypothesis of a state-dependent redistribution of DHA that may explain the observed lower circulating DHA-containing PCs in more active children through PA-induced uptake of these PCs from circulation into muscle.

Beyond individual lipid metabolites, we identified robust associations of the ratio Val/LPC a C18:1 with multiple PA measures. Given its reported associations with insulin resistance and prevalent and incident type 2 diabetes in adults^48^, this ratio is of particular interest in the context of PA and metabolic health. The lower Val/LPC a C18:1 ratios in more active children in our study may reflect better insulin sensitivity. Notably, neither valine nor LPC a C18:1 alone was associated with PA, suggesting that their ratio captures a combined metabolic signal from BCAA and PC metabolism that is not apparent in either metabolite individually. Val/LPC a C18:1 may therefore be a potential marker for capturing metabolic health that is modifiable through PA in children.

### Metabolite association patterns vary by physical activity intensity

The overlap in metabolite associations across the five PA variables broadly reflected the correlations between the corresponding activity measures, indicating that similar PA intensities are accompanied by similar metabolic profiles. Sedentary behavior and total PA showed substantial overlap, with shared metabolite associations consistently displaying opposite effect directions, as reported previously^18,21,25^. Conversely, the relatively small overlap between the associations for moderate to very vigorous PA, capturing exercise-related activity, and total PA, which constitutes mostly of light activity, suggests that activities of different intensities may engage partly distinct metabolic pathways: While TCA cycle metabolites and DHA-containing PCs were associated with activity across intensity levels, acylcarnitines and amino acids were associated predominantly with higher intensity activity and LPCs predominantly with total PA.

### Metabolite associations with habitual physical activity depend on sex

Our findings extend previous evidence of sex differences in the childhood plasma metabolome^73,74^, body composition^75^, and PA patterns, with boys generally spending less time in light PA but more time in moderate-to-vigorous PA than girls^76^. We further show that associations between habitual PA and several lipid metabolites, specifically PC ae metabolites, FFAs and LPCs, also differ between boys and girls, independently of BMI. For FFAs and LPCs, more active girls generally had higher metabolite levels, whereas more active boys generally had lower levels. In adults, lipid classes have been reported to respond to exercise in a sex-specific way, both acutely, with FFAs and LPCs among the metabolite classes showing the strongest sex differences^77^, and with habitual training, where, for example, 24 weeks of moderate-intensity exercise increased circulating triglyceride species more in women than in men^64^. One possible explanation for these findings is the known sex difference in substrate use. Females rely more on lipid metabolism during both exercise and fasting, whereas males generally utilize carbohydrates to a greater extent^78^. In adults, this difference is driven in part by estrogen, which promotes lipid oxidation, and is reflected in the higher intramuscular triglyceride content and greater fatty-acid uptake capacity (FAT/CD36) in females^79^, alongside higher glycolytic capacity, lower oxidative capacity, and lower skeletal-muscle insulin sensitivity in males^80^. Because in our study, blood samples were largely collected after fasting, circulating FFA levels likely reflect the balance between adipose lipolysis and tissue uptake. Within this framework, the higher FFAs in more active girls may reflect greater lipid mobilization and turnover, whereas the lower FFAs in more active boys may be linked to stronger suppression of fasting lipolysis due to improved insulin sensitivity. However, this interpretation is largely derived from adult studies and its relevance to childhood remains uncertain.

### Metabolite associations with habitual physical activity depend on BMI

Various metabolite associations with PA, in particular those implicated in energy-related processes such as acylcarnitines, BCAAs, and the ketone body β-hydroxybutyric acid, differed depending on BMI, suggesting that habitual PA may be accompanied by different patterns of substrate utilization in children with different body weight status independent of sex. For example, among children with lower BMI, higher activity was associated with lower circulating levels of the long chain acylcarnitine CAR 18:2, whereas the model suggests no such association among children with higher BMI. Long-chain acylcarnitines are frequently higher in obesity, where they are thought to reflect impaired fatty acid utilization^52,81^. Thus, their PA-associated reduction seen only in leaner children may indicate a greater capacity to mobilize and oxidize fatty acids in response to habitual activity. The absence of this association in children with higher BMI may indicate a reduced ability to adapt lipid oxidation pathways to increasing PA. However, BMI also modified the associations of PA with free carnitine and the CPT1 ratio (free carnitine/[CAR 16:0 + CAR 18:0]), a newborn screening marker with higher values indicating lower CPT1 enzyme activity^82^. With more PA, leaner children showed a higher ratio, whereas children with high BMI showed no or opposite associations. As reduced CPT1 activity has been linked to decreased lipid oxidation in skeletal muscles of obese adults^83^, this pattern does not align with the interpretation above, and the underlying mechanisms remain unclear.

Evidence for BMI-related differences in lipid utilization for energy metabolism was also observed for β-hydroxybutyric acid. Although β-hydroxybutyric acid was not associated with sex, BMI or habitual PA in the pairwise analyses, the interaction analyses revealed that its levels tended to be higher in more active children with higher sex- and age**-**adjusted BMI and in boys but tended to be lower in more active children with lower BMI and in girls. β-Hydroxybutyric acid is a ketone body produced in the liver when dietary carbohydrate availability is low (e.g., during fasting or prolonged exercise) and energy production must rely on fatty acids. Its plasma levels typically reflect the balance between hepatic production and utilization in peripheral tissues such as skeletal muscle^84^. The positive association between PA and β-hydroxybutyric acid in children with higher BMI and in boys may indicate relatively greater ketone production or reduced utilization, whereas the inverse association in children with lower BMI and in girls may reflect lower production or more efficient peripheral uptake and utilization. The latter would be consistent with the increased capacity of exercise-trained skeletal muscle to utilize ketone bodies^85^, which could result in lower circulating concentrations despite higher metabolic flux.

BMI-dependent association patterns were also observed for phenylalanine and the BCAAs valine and leucine. Among children with higher BMI, levels of these amino acids were lower in more active children, whereas, in leaner children, the effect was in the opposite direction. Higher circulating BCAA concentrations have been consistently observed in obese children^86^ and adults^87^ and are considered to reflect alterations in BCAA catabolism linked to mitochondrial dysfunction^88,89^. Conversely, exercise has been linked to enhanced mitochondrial function and oxidative metabolism, specifically in skeletal muscle^9^. In this context, the lower amino acid levels observed with increasing PA may indicate enhanced amino acid oxidation or utilization among children with higher BMI. On the other hand, exercise enhances muscle mass and protein synthesis^58^ and higher circulating BCAA levels have repeatedly been associated with greater lean mass in adults^59–61^, which may be explained by greater BCAA release from skeletal muscle during fasting. Therefore, in children with lower BMI, the higher BCAA levels observed in more active individuals may partly reflect relatively greater muscle mass compared with their less active (lean) peers.

Broadly, these findings raise the possibility that children with higher BMI rely less on lipid oxidation and more on amino acid catabolism during PA. Although the underlying mechanisms cannot be established from our observational data, these results are consistent with the concept that obesity is associated with reduced metabolic flexibility and altered substrate partitioning, which may modify the metabolic response to habitual PA.

### Metabolic imprint of habitual physical activity increases with age

In our study, the number of PA-associated metabolites increased with age, suggesting growing PA-related metabolic effects potentially reflecting the maturation of muscle systems, including anaerobic metabolism, sympathetic nervous system activation, and ability to recruit higher-threshold motor units from lower to higher capacities over time in childhood^90–92^. Furthermore, muscle glycogen and phosphocreatine stores^90,93^ and muscle fiber size increase with age^94^. Consistent with the age-dependent effect observed in our study, a longitudinal study reported an approximately 15% increase in leucine and isoleucine levels from 11 to 18 years of age among consistently low-active individuals. In contrast, no change was observed among high-active girls, potentially suggesting a stronger metabolic impact of sedentary behavior at older ages^24^.

### Strengths and Limitations

The rich phenotypic and metabolomic data from children at different ages, including accelerometer-assessed PA, its metabolome-wide analysis approach, and the replication of results in an independent cohort are particular strengths of our study. Nonetheless, it also comes with various notable limitations. First, PA was assessed only over a limited number of days, and short-term monitoring may not fully capture long-term behavior. Moreover, wearing an accelerometer may itself influence PA during the measurement period. Second, the applied metabolomics approach primarily targeted specific pathways and therefore does not provide a comprehensive coverage of the entire metabolome. Moreover, the measurements were performed in separate batches mostly following the times of sample collection. Thus, differences in metabolite profiles between study visits may partly reflect technical variability, rather than capturing purely age-related biological changes. Despite the longitudinal design of the study, this restricts the direct interpretation of intra-individual changes. To cope with this issue, we included visit as a random effect in the linear mixed-effects models. Third, the ToMI study used for replicating our results differed in various aspects from the CHOP cohort, including methodological differences (accelerometer devices and data processing methods, and, for certain metabolite classes, metabolomics platforms) and differences in age ranges as ToMI data was on toddlers and young children while CHOP data extended into puberty, representing developmental stages with markedly different PA patterns. Replicated PA-metabolite associations across these cohorts despite these differences therefore indicates effects that are conserved across distinct developmental stages in childhood. However, as both the discovery and the replication cohort were limited to participants from European countries, generalizability of the findings to populations with different ethnic and cultural backgrounds may still be limited. Lastly, none of the metabolite associations survived FDR correction, likely reflecting the modest effect sizes expected for habitual PA and the substantial multiple-testing burden inherent to metabolomics analyses. Also, the fewer associations at 5.5 years may partly reflect the six-month gap between PA assessment and blood sampling at this visit rather than age differences. Finally, due to the association-based nature of our analyses, no causal relationship between metabolite levels and habitual PA can be inferred from our results. Consequently, individual associations should be interpreted with caution. However, the convergence of findings at the metabolite-class level, together with replication of several associations in an independent cohort and consistency with previous observational studies, suggests that the reported patterns may reflect underlying biological relationships warranting further investigation.

## Conclusion

The World Health Organization 2020 guidelines on PA and sedentary behavior for children and adolescents identified key research gaps to better inform future recommendations^95^. Our study addresses three of proposed priorities: the use of device-based measures to quantify habitual PA, the investigation of intensity-specific associations, and the evaluation of subgroup differences. Using a metabolome-wide approach with accelerometer-based PA assessment, we identified and replicated several metabolite signatures associated with habitual PA in children, suggesting that PA is reflected in the plasma metabolome already early in life. These associations varied by PA intensity and differed by age, sex, and BMI. Future studies in larger, longitudinal cohorts integrating health outcomes are needed to further investigate the clinical relevance of these findings and to inform effective prevention strategies early in life.

## Author contributions

Conceptualization, LV and GK; formal analysis, LV; investigation, LV, JH, JF and KG; data curation, LV, JF, KG, FG, HD, WRM, VL, VG and GK; resources, HD, MA, EV, DG, AX, JE, BK, VL and VG; visualization, LV and GK; funding acquisition, JE, BK, VL, VG, and GK; supervision, GK; writing – original draft, LV and GK; writing – review & editing, all authors.

## Acknowledgements

This project has received funding from the European Union’s Horizon 2020 Research and Innovation programme under the ERA-NET Cofound action Number 727565, JPI-HDHL-INTIMIC 2021; Instituto de Salud Carlos III (ISCIII) and Next Generation EU funds for actions of the Recovery, Transformation and Resilience Plan (PRTR) under grant agreement AC21_2/00010 (IISPV); Bundesministerium für Forschung, Technologie und Raumfahrt (BMFTR) under grant agreements 01EA2203A (LMU) and 01EA2203B (HMGU); and Österreichische Forschungsförderungsgesellschaft mbH (FFG) and the MissionERA program under grant agreement FO999890543 (MUW).

The Childhood Obesity Project was funded by the 5th Framework Program [QLRTe2001e00389 & QLK1-CT-2002-30.582], the 6th Framework Program (contract number FOOD-CT-2005-007036), the 7th Framework Program (FP7-KBBE-2007-1, ref. n° 212.652; and FP7-289346-EarlyNutrition) and the European Union’s Horizon 2020 research and innovation programme under the ERA-NET Cofound action (no 727565) - JPI Call PREPHOBES (PCI2020-120.697-2, EndObesity Project) of the European Commission. This manuscript does not necessarily reflect the views of the Commission and in no way anticipates the future policy in this area. The sponsor of <u>ToMI study</u> is Societé de Produits Nestlé S.A., Switzerland (Avenue Nestlé 55, CH-1800 Vevey) – formerly named Nestec.

BK is the Else Kröner Senior professor of Paediatrics co-funded by the Else Kröner-Fresenius-Foundation, Bad Homburg. Germany, LMU and LMU University Hospitals, Munich, Germany, [No. 2020_EKSP.119. 2020]. VL holds a Serra Húnter Fellowship from Generalitat de Catalunya. GK received funding (through her institution) from the National Institutes of Health/National Institute on Aging through following grants and supplements thereto: U01AG061359 and U19AG063744. The work of VG, FG and BK is/was financially supported in part by the European Commission H2020 Programme Lifecycle-733206, the European Joint Programming Initiative Project NutriPROGRAM, the German Ministry of Education and Research, Berlin (Grant Nr. 01 GI 0825), the German Research Council (Ko912/12–1 and INST 409/224-1 FUGG); German Federal Ministry of Education and Research, Berlin, N° 01EA2101 (EndObesity) under the umbrella of the European Joint Programming Initiative “A Healthy Diet for a Healthy Life” (JPI HDHL); German Ministry of Education and Research as part of the German Center for Child and Adolescent Health (DZKJ), 01GL2406A.

The authors thank Zorica Stijepic, Stephanie-Katrin Magdalena Winterstetter and Lukas Eckerlein for HPLC-ESI-QTRAP-MS/MS and FIA-MS/MS analysis and quantitative data evaluation of ToMI study samples. In addition, we want to thank the previous laboratory staff members Olaf Uhl, Christian Hellmuth, Wolfgang Preissner, Franca Kirchberger, and all others, who have helped generating the original CHOP study datasets. Furthermore, we thank Martin Klingenspor and Markus List for their helpful advice on the analyses and their interpretation.

## Data availability

Data can be made available upon request for specific purposes and after establishing a data transfer agreement. Sharing of individual participant data was not included in the informed consent of the studies. Therefore, we are unable to place data in a public repository.

## Declaration of generative AI and AI-assisted technologies in the manuscript preparation process

During the preparation of this work, the authors used ChatGPT (OpenAI) and Claude (Anthropic) for language editing, rephrasing, and consistency checks. After using these tools, the authors reviewed and edited the content as needed and take full responsibility for the content of the manuscript.

