## Supplementary Methods and Figures 1-6 for "Metabolomic Signatures of Habitual Physical Activity in Childhood"

#### Study population

The CHOP study[1,2] (NCT00338689) was initiated as a randomized controlled trial, designed to investigate the effects of different dairy protein levels in infant formula on body composition later in life. Between October 2002 and July 2004, 1,678 healthy newborns aged ≤8 weeks were recruited across five European countries (Belgium, Germany, Italy, Poland, and Spain). They were randomized to receive an infant milk formula with either higher or lower protein content or served as controls (exclusively breastfed) until the age of 12 months. Local ethics committees of each study center approved the trial: Belgium (Comité d'Ethique de L'Hopital Universitaire des Enfants Reine Fabiola; No. CEH 14/02), Germany (Bayerische Landesärztekammer Ethik-Kommission; No. 02070), Italy (Azienda Ospedaliera San Paolo Comitato Etico; No 14/2002), Poland (Instytut Pomnik-Centrum Zdrowia Dziecka Komitet Etyczny; No 243/KE/2001), and Spain (Comité ético de investigación clínica del Hospital Universitario de Tarragona Joan XXIII). After the intervention, the children underwent multiple follow-up assessments, including assessments at 5.5, 8, and 11 years.

The ToMI study[3] (NCT02907502) was a randomized controlled trial designed after CHOP, to study the role of milk protein intake during the second year of life on child growth and later obesity risk. At ~12 months of age 1,627 children were recruited in Munich, Germany, and Tarragona/Reus, Spain, and 1,624 received an isocaloric young child milk formula with either higher or lower protein content, which they were instructed to consume during the second year of life. Ethics approval was obtained from the ethical committees of the university hospitals at the Ludwig-Maximilian University in Munich, Germany (Project Nr. 555–15) and at the Institut de Recerca Biomèdica Catalunya Sud (formerly named Institut d'Investigació Sanitaria Pere Virgili), Reus, Spain (Ref. CEIm IISPV 013/2016). Children were followed up until 6 years of age within the BiomarKid project.

#### Metabolite profiling

Metabolite profiling of plasma samples (CHOP and ToMI, from all study centers) using tandem mass spectrometry coupled with liquid chromatography (LC-MS/MS) was conducted at the MS-Core Facility of the LMU Hospital Munich (Dr. von Hauner Children's Hospital). Following the longitudinal study design, CHOP samples were measured after completion of each study visit. For ToMI, 2- and 6-year samples were collected and analyzed together in one sequence. For both, CHOP and ToMI, quality control (QC) samples were co-analyzed in six wells of each 96-well plate batch. For CHOP, adult human plasma was used as QC, whereas for ToMI, a pooled sample comprising plasma from 2- and 6-year samples was used as QC. For both studies, two commercial adult human control plasma samples (CP-I and CP-II; Recipe) were also co-analyzed in duplicate on each analytical well plate. An extended description of calibration standards, internal standards, MS-parameters, Q1/Q3 transition, chromatographic parameters and quantitation can be found in the Supporting Information of Hao et al.[4].

*CHOP sample preparation:* The frozen plasma samples were thawed to room temperature, vortexed and centrifuged for 1-2 minutes to obtain homogeneous sample solutions. Into each well of a PTFE-filter 96-well plate, attached to a 96-deep-well plate with tape, 450 µL of internal standard mixture in methanol was added using a stepper pipette, followed by the manual addition of 50 µL plasma. This plate assembly was covered with a PCR-foil, shaken for 20 min at 900 rpm at 25 °C, incubated at -20 °C for 20 min and then centrifuged at 1200 rpm for 10 min at 22 °C. The 96-well plate with the clear metabolic extract solution was covered with a PCR-foil and stored at -20 °C before LC-MS/MS sample preparation.

*ToMI sample preparation:* The frozen plasma samples were thawed at 4 °C, vortexed and centrifuged for 1-2 minutes to obtain homogeneous sample solutions. Into each well of a 96-deep-well plate, 450 µL of internal standard mixture in methanol was added using the epMotion 96XL pipettor (Eppendorf; Hamburg, Germany). Subsequently, 50 µL of plasma was added to the methanolic solution, followed by 20 cycles of aspiration and dispensing using the epMotion 96XL to precipitate the plasma proteins. The 96-well plate was covered with a PCR-foil, shaken for 20 min at 700 rpm and 25 °C, and incubated at 4 °C for 20 min. The methanolic protein slurry was manually transferred with a 12-channel pipette to a PTFE-filter plate with 96-deep-well plate assembly. The plate was covered with a PCR-foil and centrifuged at 1200 rpm for 10 min at 22 °C. The clear metabolite extract was stored at -30 °C before LC-MS/MS sample preparation[4].

*HPLC-ESI-QTRAP-MS/MS:* For targeted amino acid analysis, 50 µL of metabolite extract was esterified with n-butanol in 3 M HCl. The dried derivatives were reconstituted in 100 µL of 80:20:0.1 (v/v/v) water, methanol and formic acid and analyzed on a Waters XBridge C18 column (2.1 mm × 150 mm; 3.5 µm) using heptafluorobutyric acid as an ion-pair reagent in water and methanol. Analysis was performed on an Agilent 1100 HPLC system equipped with a binary pump, degasser, well-plate autosampler and column oven (Agilent; Waldbronn, Germany), which was coupled to an API-2000 tandem-triple-quadrupole MS/MS system (Sciex; Concord, Canada). For CHOP samples, the protocol described by Harder et al.[5] was used, whereas for ToMI samples, the Hao et al.[4,6] protocol was applied, using an 8-point calibration curve with an extended calibration range up to 1000 µmol/L. Analysis was performed in positive ionization mode.

For targeted organic acid analysis, including α-aminoadipic acid and taurine, in negative ionization mode, 100 µL of metabolite extract was dried and reconstituted in 50 µL of water containing 0.1% formic acid. Analysis was performed on a Kinetex PFP column (150 mm × 2.1 mm; 2.6 µm), using an Agilent 1260 HPLC system with SL binary pump, 2-well plate autosampler (or 8-well plate Multisampler for ToMI), and MayLab column oven, coupled to a hybrid QTRAP 4000 MS/MS system (Sciex; Ontario, Canada). Different protocols were used for CHOP[7] and ToMI[4]. For ToMI an extended 10-point calibration curve was used.

Flow-injection-analysis (FIA) of phospholipids (phosphatidylcholines (PCs), lysophosphatidylcholines (LPCs), sphingomyelins (SMs)), acylcarnitines and hexoses in CHOP samples was performed using the Agilent 1260 HPLC system with QTRAP 4000 MS/MS. A 30 µL aliquot of diluted metabolic extract was injected, prepared by diluting 9 µL of extract in 150 µL of 0.25 mM ammonium acetate in methanol. For ToMI samples, FIA-ESI-QTRAP-MS/MS analysis was performed only for PCs, LPCs, and SMs. In both studies, one-point calibration was employed for quantitation[4,8].

For targeted acylcarnitine analysis of ToMI samples, 50 µL of metabolite extract was first derivatized with n-butanol in 3 M HCl. The dried extract was then reconstituted in 50 µL methanol. Analysis of ToMI samples was performed using one-point calibration on a Kinetex EVO C18 column (150 mm × 2.1 mm; 2.6 µm) with the Agilent 1260 HPLC system coupled to a QTRAP 4000 MS/MS system in positive ionization mode[4,9,10].

In CHOP, homocysteine and cysteine were analyzed in positive ESI mode on a silica-based Cogent Diamond Hydride column (MicroSolv Technology Corporation; 150 mm × 2.1 mm; 4 µm) using the Agilent 1260 HPLC system coupled to the QTRAP 4000 MS/MS (Sciex)[11].

Free fatty acids (FFAs) in CHOP samples were analyzed using the same LC-MS/MS system on a Pursuit UPS Diphenyl column (100 mm × 3.0 mm; 1.9 µm) in negative ionization mode[12].

**Metabolomics data preprocessing and harmonization**

In CHOP, QC was performed separately for each visit (5.5, 8, and 11 years) as different QC samples were used. First, we performed a Principal Component Analysis (PCA) for each metabolite class-specific platform (using log2-transformed concentrations and imputing missing values with metabolite means) and visually inspected the score plots to identify sample outliers. Based on this step, we excluded one sample from the FIA-MS/MS data set, for which > 40% of the metabolite values were missing. Additionally, outlying QC samples were excluded based on extreme deviations in the distribution of multiple metabolites (two from organic acids and one from FIA-MS/MS data sets for 8 years). Subsequently, extreme metabolite values in QC samples were removed using a mean ± 3×SD threshold. QC samples were then used for technical batch correction. Specifically, metabolite values for each sample were divided by the mean metabolite value of QC samples within the batch in which that sample was analyzed and then multiplied with the mean for this metabolite in QC samples across all batches. For batches without available metabolite values for QC samples, batch correction was performed using the same approach but replacing QC samples with study samples. After batch correction, the quality of metabolite measurements was assessed by determining the remaining technical variation in CP-II and dispersion in QC samples. Prior to this assessment, extreme CP-II metabolite values were removed (< Q1-1.5*IQR or > Q3+1.5*IQR). Metabolites were excluded if they met the following criteria: (i) a coefficient of variation > 30% in CP-II samples (or in QC samples for acylcarnitines and phospholipids), (ii) a dispersion ratio, defined as the standard deviation of QC samples divided by that of study

samples, greater than 50%. Samples not measured across all analytical platforms were excluded to ensure dataset consistency and avoid high missingness per sample. Finally, metabolites with missingness exceeding 30% were excluded. This left 272 metabolites for 5.5 years, 258 for 8 years, and 292 for 11 years, with 233 metabolites in common across all three age groups.
For this study, we further harmonized the metabolite measurements, accounting for visit-specific differences. For this purpose, we divided metabolite values by the mean CP-II value within the corresponding visit and multiplied them with the overall mean CP-II across all visits (5.5, 8, and 11 years). This visit-level correction was not applied for acylcarnitines and phospholipids, as CP measurements were already used for calibration. Finally, missing values were imputed separately for each visit using a random forest approach (missForest::missForest, version 1.5), with a maximum of five iterations (maxiter = 5). Across all visits, the final data set included 24 amino acids (including homocysteine and cysteine (on 2 different platforms)), 18 organic acids, 50 free fatty acids (FFAs), hexoses, 26 acylcarnitines (including free carnitine), 28 LPCs, 106 PCs, 37 SMs, as well as 25 unidentified lipid metabolites labelled in this study by their *m/z*.

In ToMI, QC was performed together for samples at 1, 2, and 6 years, as QC and CP samples were the same across age visits. The QC pipeline was similar to that for CHOP. First, one study sample with high missingness (> 40%) and an outlier position in the PCA plot was removed from the FIA-MS/MS data set. There were no outlying QC samples in ToMI. We did not remove any values beyond the 3 SD threshold before batch correction. Otherwise, batch correction was conducted as described for the CHOP pipeline but using QC medians instead of means. Quality of measured metabolites was assessed based on the variation of CP sample values and dispersion ratio using the QC samples after removal of values < Q1-3*IQR or > Q3+3*IQR from CP and QC sample measurements. Metabolites were excluded if coefficients of variation of CP-I and CP-II were higher than 30% and the dispersion ratio was greater than 50%. Samples not measured across all analytical platforms were excluded. Lastly, metabolites with missingness > 30% were excluded and remaining missing values were imputed using the same approach as for CHOP. The final ToMI metabolomics dataset included 22 amino acids, 15 organic acids, 60 acylcarnitines (including free carnitine), and 167 phospholipids (91 PCs, 25 LPCs, and 51 SMs).

12. Hellmuth C, Weber M, Koletzko B, Peissner W. Nonesterified fatty acid determination for functional lipidomics: comprehensive ultrahigh performance liquid chromatography-

tandem mass spectrometry quantitation, qualification, and parameter prediction. *Anal Chem*. 2012;84(3):1483-1490. doi:10.1021/ac202602u

### Supplementary Figures

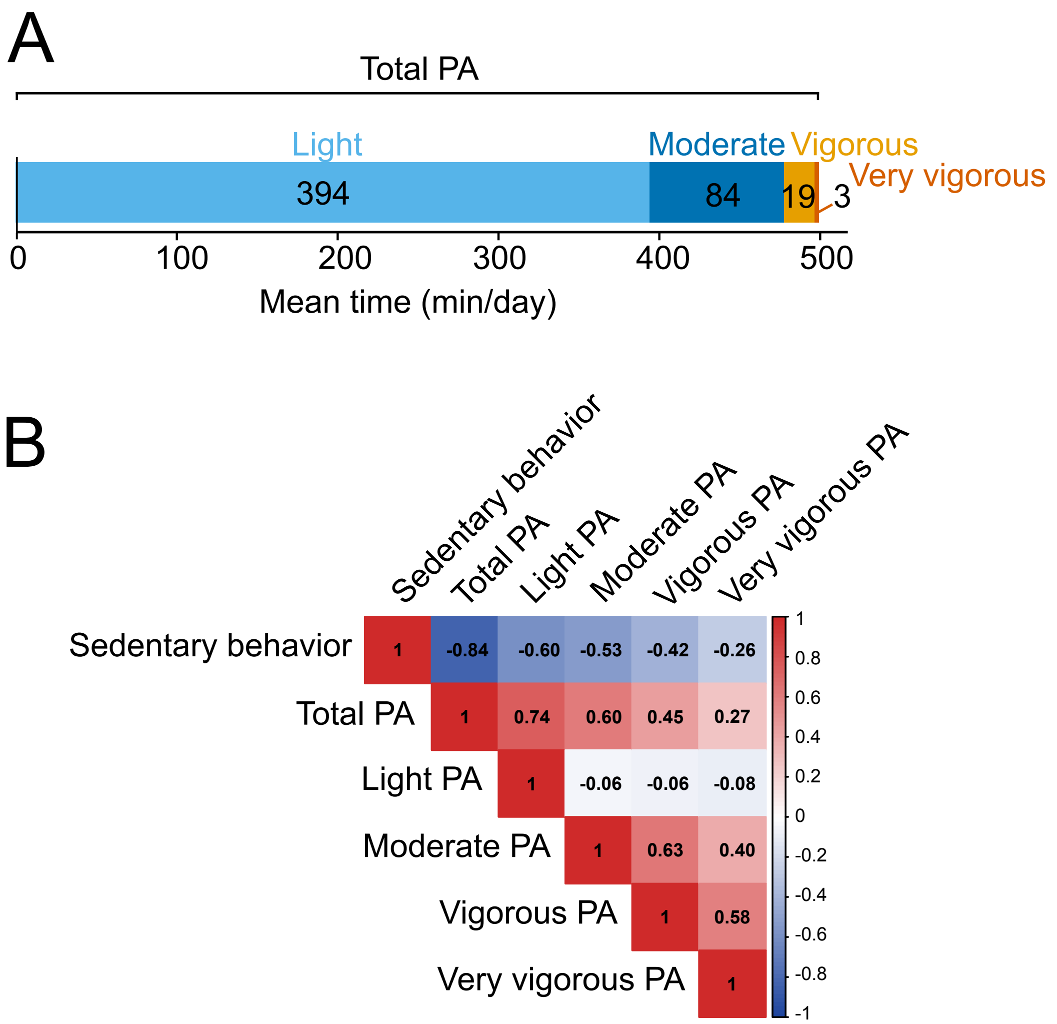


**Supplementary Figure 1: Physical activity (PA) variables in CHOP at 5.5, 8, and 11 years. A)** Mean daily time spent in each PA intensity and its contribution to total PA. **B)** Heatmap of Spearman correlation matrix for PA measures. The heatmap depicts Spearman rank correlation coefficients between each pair of six different PA measures. Vigorous and very vigorous PA were coded as ordered tertiles (1, 2, 3). The color scale indicates the strength of the correlations, with darker blue representing strong negative relationships and darker red stronger positive correlations.

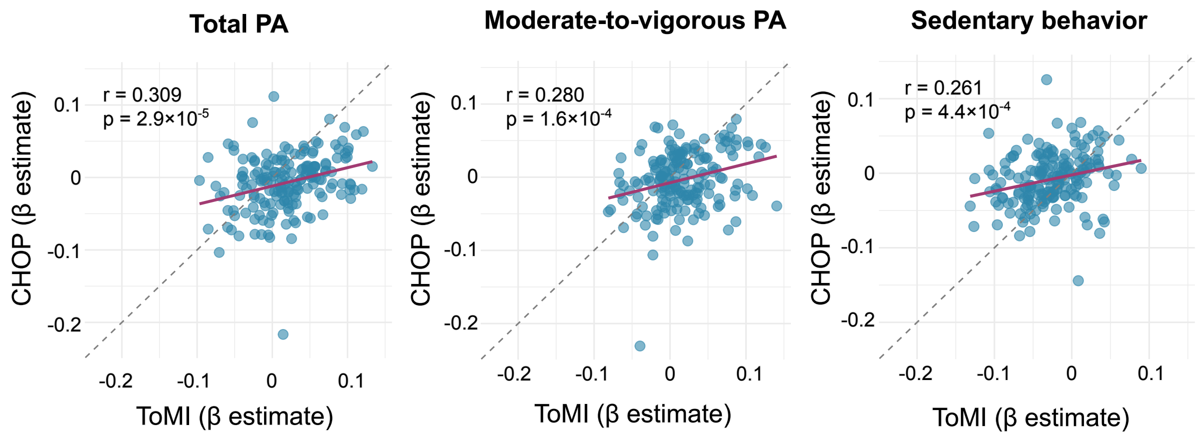


**Supplementary Figure 2: Correlation of metabolite-physical activity (PA) associations between ToMI and CHOP.** Scatter plots show β effect estimates for PA variables from linear mixed models in ToMI (x-axis) and CHOP (y-axis). Each point represents β effect value of PA for one metabolite. Solid red lines show the fitted linear regression between estimates, and dashed lines indicate the line of equality (y = x). Pearson correlation coefficients (r) and corresponding p values are displayed for each PA variable. Metabolite ratios and sums were excluded from this analysis.

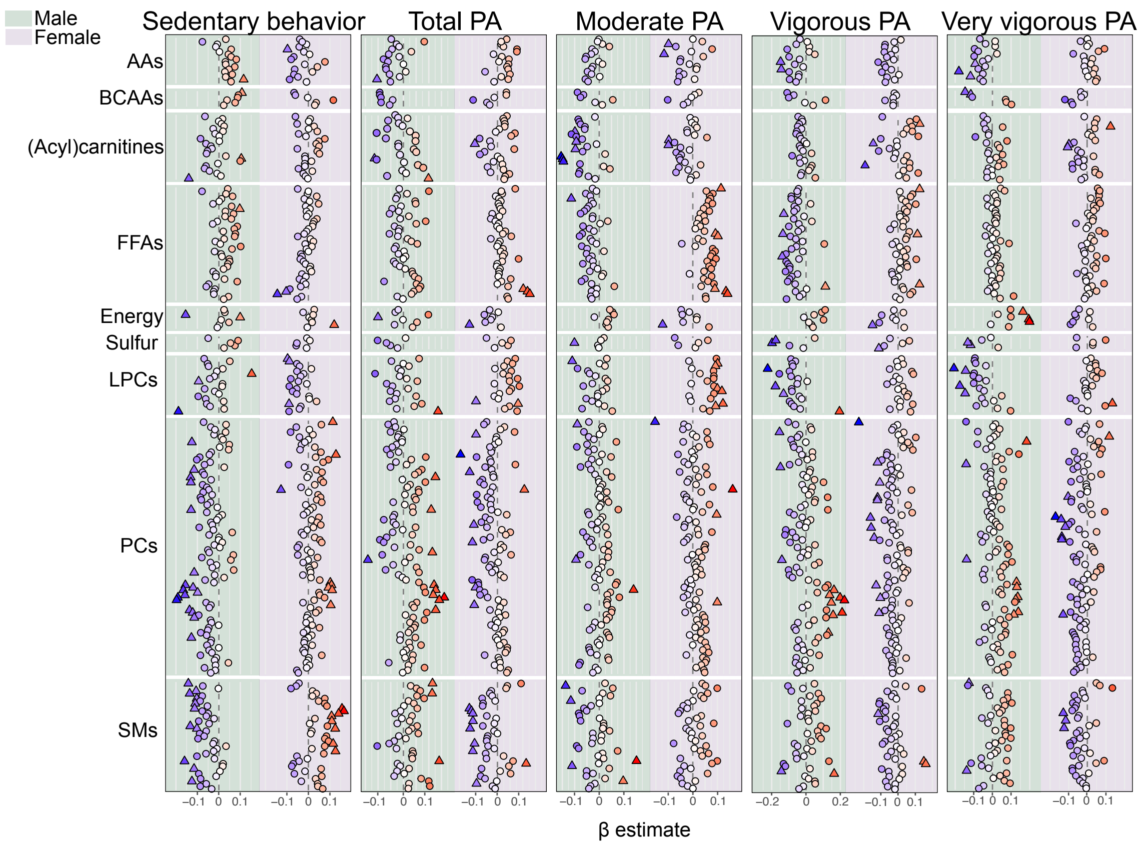


**Supplementary Figure 3: Sex-stratified analysis of associations between metabolites and physical activity (PA) variables in CHOP.** Color represents β estimates, with darker blue reflecting stronger negative and darker red stronger positive associations. Significant associations ($p < 0.05$) are depicted as triangles while not significant metabolites are depicted as circles. Metabolite ratios and sums are not included in the plot. AA: amino acids; BCAA: branched-chain amino acids; Energy: metabolites involved in central energy metabolism (e.g., TCA cycle metabolites, glucose); FFAs: free fatty acids; LPCs: lysophosphatidylcholines; PCs: phosphatidylcholines; SMs: sphingomyelins; Sulfur: metabolites involved in sulfur metabolism (e.g., cysteine, homocysteine and taurine).

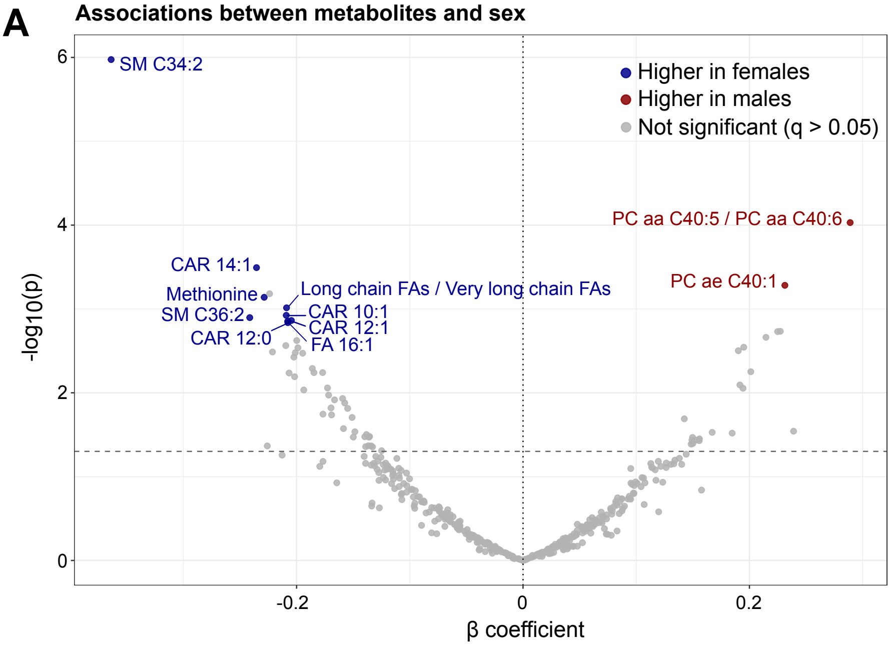


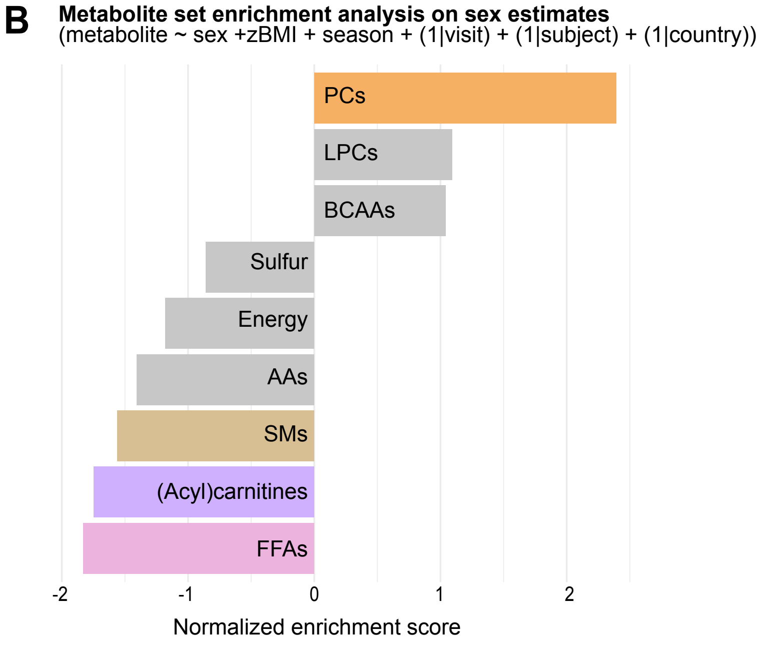


**Supplementary Figure 4: Associations between metabolites and sex in the CHOP. A)** Volcano plot showing associations between metabolites and sex. Each point represents one metabolite variable. Metabolites significantly associated with sex after controlling for the false discovery rate using the Benjamini–Hochberg procedure ($q < 0.05$) are highlighted in red or blue, while non-significant metabolites are shown in grey. Metabolite sums and ratios were considered significant only if they met both criteria of $q < 0.05$ and a p-gain > 1. Metabolite variables with higher levels in boys are shown in red, and metabolites with lower levels in boys are shown in blue. **B)** Metabolite set enrichment analysis based on β effect estimates from the association models shown in panel A. Colored bars show metabolite groups significantly enriched after multiple-testing correction ($q < 0.05$); grey bars denote non-significant groups. A positive normalized enrichment score (NES) indicates higher levels of that metabolite group in boys relative to girls; a negative NES indicates lower levels in boys relative to girls. AA: amino acids; BCAA: branched-chain amino acids; Energy: metabolites involved in central energy metabolism (e.g., TCA cycle metabolites, glucose); FFAs: free fatty acids; LPCs: lysophosphatidylcholines; PCs: phosphatidylcholines; SMs: sphingomyelins; Sulfur: metabolites involved in sulfur metabolism (e.g., cysteine, homocysteine, and taurine).

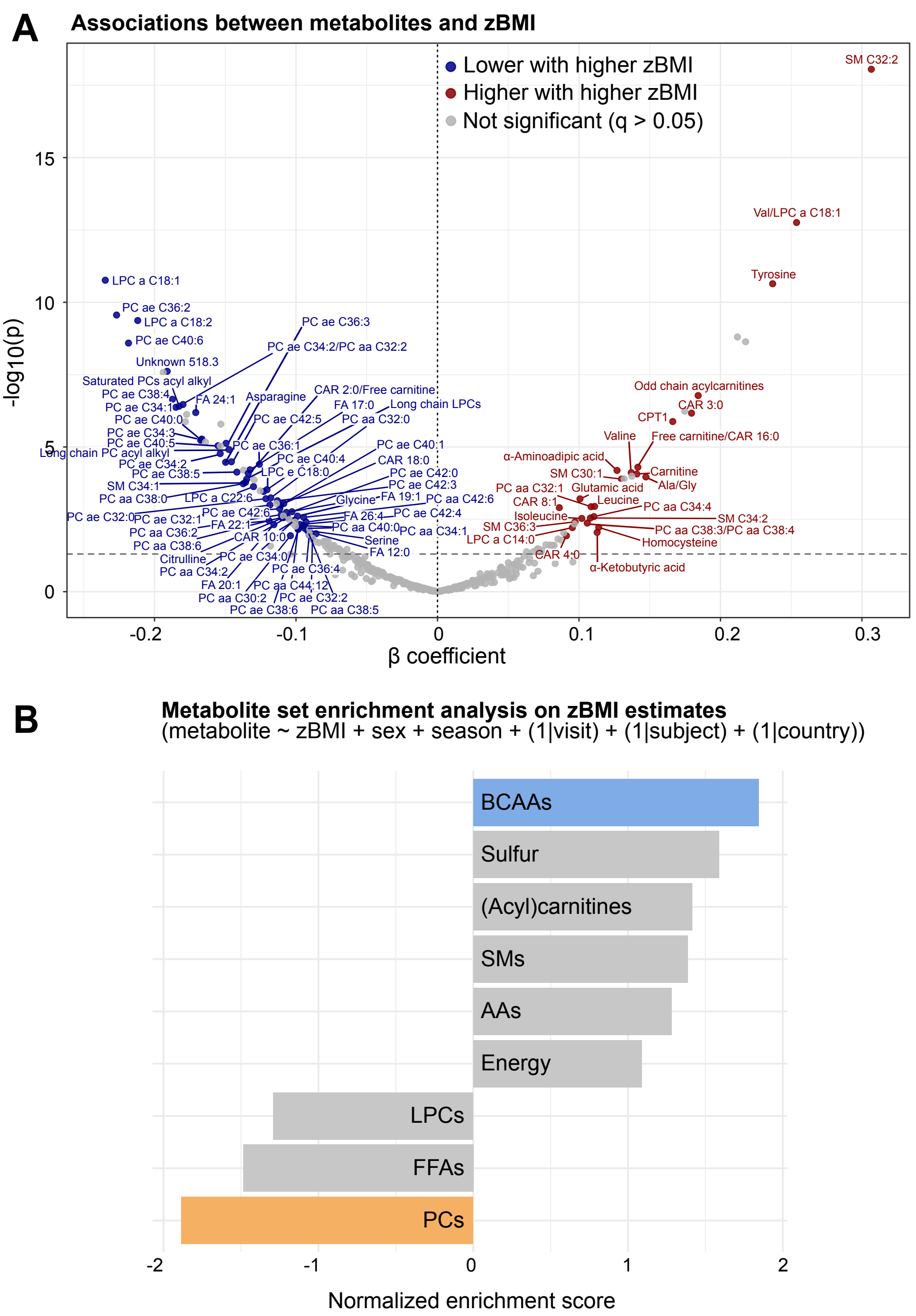


**Supplementary Figure 5: Associations between metabolites and zBMI in the CHOP. A)** Volcano plot showing associations between metabolites and zBMI. Metabolites significantly associated with zBMI after controlling for the false discovery rate using the Benjamini–Hochberg procedure ($q < 0.05$) are highlighted in red (higher metabolite levels associated with higher zBMI) or blue (lower metabolite levels associated with higher BMI), while non-significant metabolites are shown in grey. Metabolite sums and ratios were considered significant only if they met both criteria of $q < 0.05$ and a p-gain > 1. **B)** Metabolite set enrichment analysis based on β effect estimates from the association models shown in panel A. Colored bars show metabolite groups significantly enriched after multiple-testing correction ($q < 0.05$); grey bars denote non-significant groups. A positive normalized enrichment score (NES) indicates that the metabolite group is enriched with metabolites showing higher levels at higher zBMI; a negative NES indicates enrichment of metabolites with lower levels at higher zBMI. AA: amino acids; BCAA: branched-chain amino acids; Energy: metabolites involved in central energy metabolism (e.g., TCA cycle metabolites, glucose); FFAs: free fatty acids; LPCs: lysophosphatidylcholines; PCs: phosphatidylcholines; SMs: sphingomyelins; Sulfur: metabolites involved in sulfur metabolism (e.g., cysteine, homocysteine, and taurine).

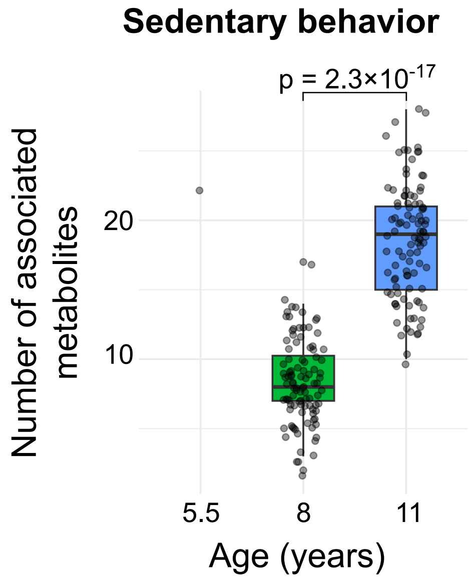


**Supplementary Figure 6: Number of metabolites associated with sedentary behavior at 5.5, 8, and 11 years in CHOP.** To ensure comparability across ages, samples at 8 and 11 years were randomly down-sampled to match the sample size at 5.5 years and this procedure was repeated 100 times. Linear mixed-effects models were fitted separately for each age and each down-sampling iteration. The figure shows the number of significantly associated metabolites obtained in each of the 100 iterations for ages 8 and 11 years. At 5.5 years, no down-sampling was required and the model was fitted only once.
